# Classifying chronic pain using multidimensional pain-agnostic symptom assessments and clustering analysis

**DOI:** 10.1101/2021.04.21.21255885

**Authors:** Gadi Gilam, Eric M. Cramer, Kenneth A. Webber, Maisa S. Ziadni, Ming-Chih Kao, Sean C. Mackey

## Abstract

Chronic pain conditions present in various forms, yet all feature symptomatic impairments in physical, mental, and social domains. Rather than assessing symptoms as manifestations of illness, we used them to develop a chronic pain classification system. A cohort of real-world treatment-seeking patients completed a multidimensional patient-reported registry as part of a routine initial evaluation in a multidisciplinary academic pain clinic. We applied hierarchical clustering on a training subset of 11448 patients using nine pain-agnostic symptoms. We then validated a three-cluster solution reflecting a graded scale of severity across all symptoms and eight independent pain-specific measures in additional subsets of 3817 and 1273 patients. Negative affect-related factors were key determinants of cluster assignment. The smallest subset included follow-up assessments that were predicted based on baseline cluster assignment. Findings provide a cost-effective classification system that promises to improve clinical care and alleviate suffering by providing putative markers for personalized diagnosis and prognosis.

## Introduction

Chronic pain is a global epidemic reflecting a health care crisis for the person suffering from it, their family, and society as a whole (*1*–*3*). More than 100 million individuals are impacted by various chronic pain conditions in the US alone, with medical expenses and lost productivity costing more than $635 billion annually and projected to become much worse (*4*–*6*). Primary chronic pain conditions present in various shapes and forms, commonly classified by anatomical location of experienced pain, from low-back pain and headaches to pelvic or bladder pain, including wide spread non-specific or overlapping pain (*7*). However, common to all conditions is a global functional impairment that is manifested in the experience of multiple physical, “mental”, and social health symptoms, reflective of the biopsychosocial model of shared etiological factors across chronic pain conditions (*7*–*10*). While various studies aimed to uncover and classify sub-groups of chronic pain (*11*–*19*), little is known whether a combination of domain-general symptoms agnostic to pain can be used to classify one’s chronic pain condition, and subsequently serve as potential markers for clinical diagnosis and prognosis (*20, 21*). A symptom-based approach may also reveal potentially modifiable factors as targets for therapeutic interventions. Therefore, we suggest a reversal of the paradigm – instead of assessing patient-reported symptoms as features of the a-priori determined pain condition, we examined whether such symptoms may serve to classify current and predict future pain condition. If confirmed, our approach could be used to support personalized and efficient treatment of individuals with chronic pain.

We implemented unsupervised machine learning, specifically agglomerative hierarchical clustering analysis (*22*–*24*), on multidimensional patient-reported symptoms that assess physical, mental, and social health status factors, to identify idiosyncratic groups, or clusters of patients with chronic pain. Patients reflected a real-world clinical population with a heterogeneous mix of pain conditions seeking treatment at a tertiary academic pain clinic. As part of their routine initial evaluation, they completed multidimensional patient-reported assessments using Stanford’s CHOIR registry-based learning health system (Figure 1A) (*25, 26*). We used nine symptoms for clustering based on the National Institute of Health’s (NIH) Patient-Reported Outcomes Measurement Information System (PROMIS), which was designed and validated for precise and efficient measurement of health-related symptoms in patients with a wide variety of chronic health conditions (*27*). These symptoms were agnostic to nine pain-specific measures that we subsequently used to validate the diagnostic-like nature of the data-driven clusters independently.

**Fig. 1.**
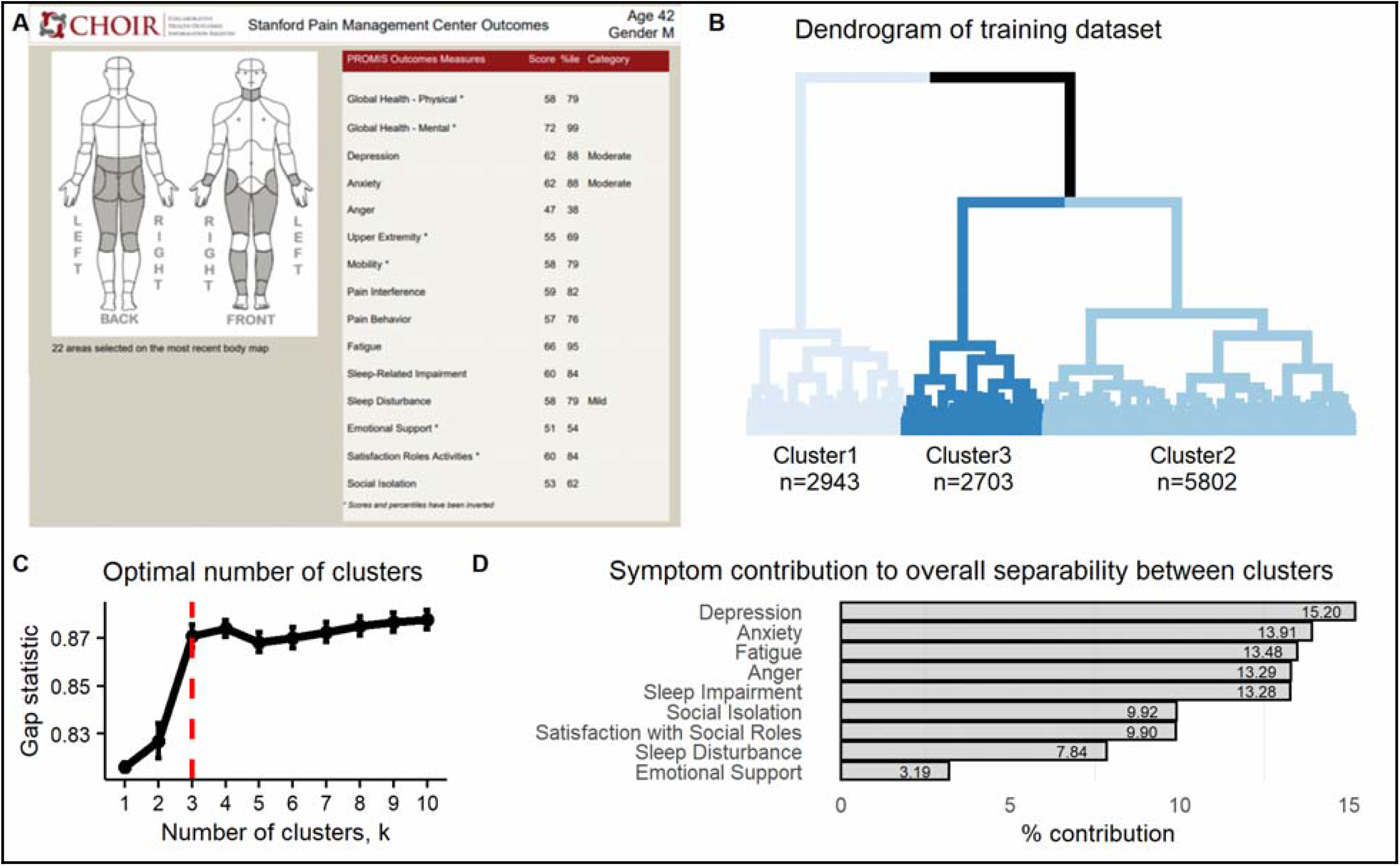
Cluster development. (**A**) An illustration of a simulated CHOIR report used at the Stanford Pain Clinics. The CHOIR body map with marked regions in pain is on the left, and multiple normalized symptom scores are listed on the right. (**B**) The dendrogram reflecting results of the agglomerative hierarchical clustering algorithm as implemented on the training dataset (n=11448) and using the nine clustering symptoms. The three-cluster solution is reflected by the different shades of blue per cluster. Cluster1 comprised of 25.71% of the patients (n=2943), Cluster2 of 50.68% (n=5802), and Cluster3 of 23.61% (n=2703). (**C**) The plot shows the gap statistic values for different k number of clusters, and indicating with a red dashed line the optimal solution of k=3 since it is the smallest value of k that is within one standard deviation of the value of k that maximizes the gap statistic. The error bars represent one standard error of the estimated gap statistic. (**D**) The plot shows the percent contribution to the overall separability between clusters of each of the nine clustering symptoms, in order from most contributing (Depression=15.20%) to least (Emotional Support=3.19%).

Mechanistically, we aimed to uncover whether the multivariate pattern of symptoms and pain-specific measures characterizing each identified cluster reflects a general graded scale of severity or a differential pattern. Furthermore, given the centrality and comorbidity of mental health related factors with chronic pain, predominantly negative affect-related symptoms such as anxiety, depression, and anger (*9, 28, 29*), we expected these symptoms to be key drivers for the determination of cluster assignment, thus highlighting them as targets for treatment. We based cluster discovery on a training dataset of 11448 patients and subsequently validated it in two additional datasets of 3817 and 1273 patients. The later dataset included follow-up assessments allowing us to examine if cluster assignment at baseline would be predictive of pain-related measures at follow-up, thus providing potential prognostic-like validation of the identified clusters. Finally, we examined the dynamics across assigned clusters between time-points.

## Materials and Methods

### General data acquisition procedures and dataset definition

Data were collected using Stanford University’s CHOIR (http://choir.stanford.edu), a registry-based, learning health care system that administers an electronic survey assessing self-reported demographic information, medical history, and multiple domains of health status in real-world clinical settings (Figure 1A) (*30*). Patients presenting for consultation at Stanford Pain Management Center locations throughout the San Francisco Bay Area and broader Norther California region, USA, with the main site located in Redwood City, complete the survey as part of their routine clinical care. While intended for completion at home using personal computers or hand-held devices, patients may complete the survey before their appointment at clinic check-in using a tablet computer. Survey completion is encouraged, yet optional, and based on patients’ willingness and ability to collaborate. Patients may therefore choose not to respond to certain items or assessments. These procedures were approved by the Stanford University School of Medicine Institutional Review Board (IRB). Informed consent was waived by the IRB, as CHOIR data were collected for clinical care and quality improvement purposes.

Data analyzed were from a retrospective review of all collected surveys since CHOIR’s inception in October 2013 through August 2019. Our initial data extraction included 24389 records, from which we removed records based on the following criteria: non-completed or test records (6002), missing data in any of the nine measures used for clustering (as detailed below; 1651), duplicated records (136), and age below 18 years (62). From the resulting 16538 surveys belonging to 16538 different patients, we extracted a *longitudinal* dataset of 1273 patients with a follow-up survey between 3-12 months later, and again with a minimal requirement of having complete data for the nine assessments used for clustering at both time points. We chose this time frame since 3 months is considered the minimal threshold for diagnosing primary chronic pain (*7*). The upper threshold of 12 months allowed to keep a substantially large proportion of the dataset for cluster discovery validation. The resulting 15265 patients were randomly split based on a 75%:25% allocation into a *training* dataset of 11448 patients used for cluster discovery and an additional *validation* dataset of 3817 patients.

### Measures

#### Demographic characteristics

Demographic characteristics included age, sex, ethnicity, race, marital status, and years of education.

#### Clustering symptoms

The nine symptoms assessing health-related functionality and used as the basis for the clustering procedures were from the National Institute of Health’s (NIH’s) Patient-Reported Outcomes Measurement Information System (PROMIS) (*27, 31*–*35*). We divided these nine symptoms into three domains: (1) the physical domain (*fatigue, sleep disturbance*, and *sleep impairment)*, (2) the mental or negative affect domain (*depression, anxiety*, and *anger)*, and (3) the social domain (*social isolation, emotional support*, and *satisfaction with social roles and activities)*. Response items are contextualized to the frequency of the experienced symptom in the past seven days (e.g., “in the past seven days how often did you feel tired?”, “in the past seven days I felt worthless”), and responses were marked on a 1-5 scale (1 = never, 5 = always). Each assessment was completed using computerized adaptive testing (CAT), based on item response theory-derived metrics. CAT reduces the time needed to complete each assessment because patients respond only to a subset of items for each PROMIS item bank, until resulting measurements meet preset criteria of below three standard errors (*27, 36*). Most patients complete 4-5 items per assessment, and take about 15 minutes to complete these nine symptoms.

Ultimately, a standardized T score for each PROMIS symptom is generated for each patient. A score of 50 reflects the mean of the US general population, with a standard deviation (SD) of 10. Higher scores reflect more of the measures’ symptom. We further extracted data of a PROMIS-based global health measure, specifically the G*lobal Health Mental* subscale that consisted of four items assessing general mental health, quality of life, satisfaction with social activities, and emotional problems (*37*). While for most measures such as fatigue or depression, higher T scores indicated a worse condition, for emotional support, satisfaction with social roles, global health mental, and physical function (see below), higher T scores reflected a better condition. Further details regarding measure development and validation are available at http://www.healthmeasures.net.

#### Pain-specific measures

Pain-specific measures were used independently of the clustering process to validate the diagnostic-like nature of the data-driven generated clusters in terms of pain-related constructs. A composite score of *pain intensity* was calculated by averaging three self-reported pain intensity measures. These measures used a common and validated (*38*) 11-point numeric rating scale of 0-10 (0=no pain, 10=pain as bad as you can imagine) for worst and average pain in the last seven days, and current pain. The *number of body segments* in which chronic pain is experienced were self-reported by patients, who were asked to mark locations of pain on a reliable and valid CHOIR body map scheme that included 36 anterior and 38 posterior symmetrical body segments for a maximum total of 74 segments (Figure 1A) (*39*). This measure was used to reflect the extent of pain throughout the body. A group of physicians recoded these 74 body segments into 11 *body regions* (Table S1, Figure S1) subsequently used to examine specific locations in which patients experienced pain. *Pain duration* was calculated as the number of months from onset of chronic pain that was self-reported by patients.

Additional measures assessed using PROMIS instrumentation included *pain interference* with daily life activities, *pain behavior*, and *physical function* (*27, 33*). In September 2016 and moving forward, physical function was assessed using two separate measures in CHOIR, reflecting physical function of the *upper extremity*, and *lower mobility* (*40*). Across the entire dataset, most patients had the two separate measures (61.32%). We analyzed each of the three physical function measures separately to be able to differentiate between them.

The last pain-specific measure was *pain catastrophizing*, reflecting maladaptive cognitions such as rumination, magnification, and helplessness, in response to actual or anticipated pain. Pain catastrophizing has been associated with poor outcomes, maintenance, and worsening of chronic pain illness (*41*–*43*). We used the Pain Catastrophizing Scale (PCS) which previously demonstrated sound psychometric properties (*44*–*46*) to measures the frequency with which a patient engages in catastrophic thought patterns, and consists of 13 self-reported items on a 0-4 scale (0=not at all, 10=all the time).

#### Statistical analysis

Programming and analyses were conducted using a combination of R version 4.0.0 (*47*), RStudio version 1.2.5042 (*48*), and IBM® SPSS® version 26. Relevant open source R codes are available at <u>TBA</u>.

#### Cluster discovery

Hierarchical clustering is a well-established unsupervised machine learning technique that aims to discover groups or clusters of observations within a dataset without needing to a-priori determine the specific characteristics of each cluster (*22*–*24*). Observations within the same cluster are expected to have similar characteristics, while different clusters are expected to have dissimilar characteristics. A cluster-tree diagram or dendrogram is a mathematical and pictorial representation of a cluster solution.

We implemented an AHCA on the training dataset and using the nine clustering symptoms, to assign each patient to a cluster. AHCA implements an iterative process in which the two most similar observations (i.e., patients, or groups of patients) are fused to form a superordinate cluster until all observations belong to one single cluster. Two parameters important for this process are a distance metric that determines how similar observations are to each other, and a linkage method to fuse similar observations. The agglomerative coefficient can then assess how tightly packed each cluster is within a cluster solution. We used the Euclidian distance metric combined with the Ward linkage method as it optimized the agglomerative coefficient compared to four other linkage methods (Table S5).

We subsequently used the gap statistic to determine the optimal number of clusters, *k* (*50*). The gap statistic compares the within-cluster sum of squares of a certain k-clusters solution to the expected within-cluster sum of squares under a null distribution with no clusters. An ideal solution will have a small within cluster sum of squares, and therefore a large gap statistic. We calculated the gap statistic for *k* between 1 and 10. The smallest value of *k* that is within one standard deviation of the value of *k* that maximizes the gap statistic should be chosen as the optimal number of clusters.

We next aimed to determine the relative importance of each clustering symptom to the clustering process, i.e. to the separability between clusters. We computed the cluster centroid (*22, 24*), which is the average value of each clustering symptom for all of the observations in that cluster, and then calculated the total Euclidian distance between all cluster centroids. The average amount each clustering symptom contributed to the distance between each clusters’ centroid, divided by the total sum of all clustering symptoms’ contribution to the total Euclidean distance between all cluster centroids, provides a percent contribution to the overall separability between clusters.

#### Cluster characterization, reliability, and validity

Univariate analysis of variance (ANOVA) and subsequent t-tests were used to examine differences in clustering symptoms and in pain-specific measures between the identified clusters, with Bonferroni correction applied to account for multiple comparisons. Chi^2^ tests were used to examine the differential distribution of demographic factors and of body regions between the clusters and determine whether specific body regions were associated with any of the identified clusters. This sequence of tests was conducted initially on the training dataset to assess the clusters’ diagnostic-like potential, and subsequently on the validation and the baseline of the longitudinal datasets, to assess the reliability of cluster assignment and validate the cluster’s characteristics in other sets of patients. A nearest centroid classifier (*22, 24*) was generated to assign or label a cluster to a “new” patient, based on the shortest Euclidian distance between the values of the clustering symptoms of that patient and each clusters’ centroids.

#### Predictive validation and cluster dynamics over time

Univariate analyses as described above were used to examine differences in clustering symptoms and in pain-specific measures between clusters as assigned at baseline, using data from the follow-up. To control for time-related effects, we added the number of days between the two assessments as covariate. This provided prognostic-like validation of the clusters.

Next, the nearest centroid classifier was implemented on the follow-up dataset to assess patient movement across clusters between the baseline and follow-up time points. Also, we used a bootstrap procedure (*23, 24*) to assess whether patients’ movement across clusters over time was due to potential error in measurement of the clustering symptoms, or potentially to the clusters’ ability to portray real improved or worsening of their condition. Since the PROMIS CAT engine uses a criterion of below three standard errors to calculate the final T score (*27, 36*), we randomly jittered the original T score for each patient and for each of the clustering symptoms at baseline, within ± three standard errors. The nearest centroid classifier was implemented on each patient’s simulated data to assign a cluster. We then assessed movement across clusters between the simulated dataset and the follow-up dataset, and calculated the number and subsequently the percent of patients moving across clusters. This procedure repeated 1000 times to generate a bootstrapped distribution of the percent of patients moving across clusters within measurement error. This distribution allowed us to calculate the probability of the actual percent of patients moving across clusters between the baseline and follow-up time points being attributed to measurement error.

## Results

### Demographic characteristics

Demographic characteristics of study participants are described in Table 1 (see also Table S1).

**Table 1.** Participants’ demographic information as per dataset and across the three clusters. Number of patients is indicated, with % in parenthesis. * Reflects the results of a Chi^2^ test (categories with less than a minimum of 5 patients per group were removed) comparing across clusters. Similar tests across datasets found no differences (*p*-values>0.74).

| N (%) | Total | Cluster1 | Cluster2 | Cluster3 | P* |
| --- | --- | --- | --- | --- | --- |
| **Training dataset** |  |  |  |  |  |
| **Total:** | 11448 (69.22) | 2943 (25.71) | 5802 (50.68) | 2703 (23.61) |  |
| **Age (years):** |  |  |  |  | 0.95 |
| 18-29 | 1398 (12.21) | 372 (12.64) | 660 (11.38) | 366 (13.54) |  |
| 30-39 | 1966 (17.17) | 510 (17.32) | 954 (16.44) | 502 (18.57) |  |
| 40-49 | 2127 (18.58) | 506 (17.19) | 1029 (17.74) | 592 (21.90) |  |
| 50-59 | 2454 (21.44) | 564 (19.16) | 1264 (21.79) | 626 (23.16) |  |
| 60-69 | 1974 (17.22) | 543 (18.45) | 1048 (18.06) | 380 (14.06) |  |
| ≥70 | 1512 (13.21) | 442 (15.02) | 836 (14.41) | 234 (8.66) |  |
| No response | 20 (0.17) | 6 (0.2) | 11 (0.19) | 3 (0.11) |  |
| **Sex** |  |  |  |  | 0.95 |
| Female | 7340 (64.12) | 1817 (61.74) | 3700 (63.77) | 1823 (67.44) |  |
| Male | 3723 (32.52) | 1025 (34.83) | 1904 (32.82) | 794 (29.37) |  |
| No response | 385 (3.36) | 101 (3.43) | 198 (3.412) | 86 (3.18) |  |
| **Ethnicity** |  |  |  |  | 0.98 |
| Hispanic/Latino | 1162 (10.15) | 304 (10.33) | 515 (8.88) | 343 (12.69) |  |
| Non-Hispanic/Non-Latino | 8527 (74.48) | 2230 (75.77) | 4401 (75.85) | 1896 (70.14) |  |
| Patient refused | 349 (3.05) | 94 (3.19) | 180 (3.1) | 75 (2.77) |  |
| Unknown | 413 (3.61) | 111 (3.77) | 211 (3.64) | 91 (3.37) |  |
| No response | 997 (8.71) | 204 (6.93) | 495 (8.53) | 298 (11.02) |  |
| **Race** |  |  |  |  | 0.99 |
| American Indian or Alaska Native | 48 (0.42) | 9 (0.31) | 22 (0.38) | 17 (0.63) |  |
| Asian | 935 (8.17) | 287 (9.75) | 469 (8.08) | 179 (6.62) |  |
| Asian, non-Hispanic | 8 (0.07) | 1 (0.03) | 5 (0.09) | 2 (0.07) |  |
| Black or African American | 405 (3.54) | 102 (3.47) | 184 (3.17) | 119 (4.4) |  |
| Black, non-Hispanic | 8 (0.07) | 3 (0.1) | 2 (0.03) | 3 (0.11) |  |
| Native American, Hispanic | 1 (0.01) | 0 (0) | 0 (0) | 1 (0.04) |  |
| Native American, non-Hispanic | 1 (0.01) | 0 (0) | 1 (0.02) | 0 (0) |  |
| Native Hawaiian or Other Pacific | 65 (0.57) | 14 (0.48) | 35 (0.6) | 16 (0.59) |  |
| Other | 1921 (16.78) | 487 (16.55) | 929 (16.01) | 505 (18.68) |  |
| Other, Hispanic | 11 (0.1) | 1 (0.03) | 9 (0.16) | 1 (0.04) |  |
| Other, non-Hispanic | 8 (0.07) | 2 (0.07) | 5 (0.09) | 1 (0.04) |  |
| Patient Refused | 326 (2.85) | 84 (2.85) | 170 (2.93) | 72 (2.66) |  |
| Unknown | 443 (3.87) | 113 (3.84) | 230 (3.96) | 100 (3.7) |  |
| White | 6148 (53.7) | 1598 (54.3) | 3186 (54.91) | 1364 (50.46) |  |
| White, Hispanic | 2 (0.02) | 1 (0.03) | 0 (0) | 1 (0.04) |  |
| White, non-Hispanic | 106 (0.93) | 30 (1.02) | 54 (0.93) | 22 (0.81) |  |
| No response | 1012 (8.84) | 211 (7.17) | 501 (8.63) | 300 (11.1) |  |
| **Marital Status** |  |  |  |  | 0.65 |
| Married | 5969 (52.14) | 1809 (61.47) | 3031 (52.24) | 1129 (41.77) |  |
| Separated | 237 (2.07) | 37 (1.26) | 111 (1.91) | 89 (3.29) |  |
| Widowed | 437 (3.82) | 99 (3.36) | 246 (4.24) | 92 (3.4) |  |
| Never Married | 2149 (18.77) | 489 (16.61) | 1065 (18.35) | 595 (22.01) |  |
| Living Together | 671 (5.86) | 157 (5.33) | 345 (5.95) | 169 (6.25) |  |
| Divorced | 1218 (10.64) | 229 (7.78) | 623 (10.73) | 366 (13.54) |  |
| No response | 767 (6.7) | 123 (4.18) | 381 (6.57) | 263 (9.73) |  |
| **Education (years):** |  |  |  |  | 0.37 |
| ≤12 | 342 (2.99) | 73 (2.48) | 159 (2.74) | 110 (4.07) |  |
| 13-16 | 3346 (29.23) | 742 (25.21) | 1606 (27.68) | 998 (36.92) |  |
| 17-20 | 6033 (52.7) | 1687 (57.32) | 3135 (54.03) | 1211 (44.80) |  |
| ≥21 | 1025 (8.95) | 327 (11.11) | 554 (9.55) | 144 (5.33) |  |
| No response | 702 (6.13) | 114 (3.87) | 348 (6) | 240 (8.88) |  |
| **Validation dataset** |  |  |  |  |  |
| **Total:** | 3817 (23.08) | 931 (24.39) | 2346 (61.46) | 540 (14.15) |  |
| **Age (years):** |  |  |  |  | 0.76 |
| 18-29 | 490 (12.84) | 119 (12.78) | 299 (12.75) | 72 (13.33) |  |
| 30-39 | 644 (16.87) | 156 (16.76) | 386 (16.45) | 102 (18.89) |  |
| 40-49 | 713 (18.68) | 144 (15.47) | 439 (18.71) | 130 (24.07) |  |
| 50-59 | 768 (20.12) | 165 (17.72) | 485 (20.67) | 118 (21.85) |  |
| 60-69 | 682 (17.81) | 198 (21.27) | 414 (17.65) | 70 (12.96) |  |
| ≥70 | 510 (13.36) | 147 (15.79) | 316 (13.47) | 47 (8.7) |  |
| No response | 10 (0.26) | 2 (0.21) | 7 (0.3) | 1 (0.19) |  |
| **Sex** |  |  |  |  | 0.87 |
| Female | 2496 (65.39) | 589 (63.27) | 1564 (66.67) | 343 (63.52) |  |
| Male | 1193 (31.25) | 322 (34.59) | 698 (29.75) | 173 (32.04) |  |
| No response | 128 (3.35) | 20 (2.15) | 84 (3.58) | 24 (4.44) |  |
| **Ethnicity** |  |  |  |  | 0.87 |
| Hispanic/Latino | 386 (10.11) | 94 (10.1) | 221 (9.42) | 71 (13.15) |  |
| Non-Hispanic/Non-Latino | 2797 (73.28) | 688 (73.9) | 1747 (74.47) | 362 (67.04) |  |
| Patient refused | 122 (3.2) | 36 (3.87) | 68 (2.9) | 18 (3.33) |  |
| Unknown | 151 (3.96) | 47 (5.05) | 87 (3.71) | 17 (3.15) |  |
| No response | 361 (9.46) | 66 (7.09) | 223 (9.51) | 72 (13.33) |  |
| **Race** |  |  |  |  | 0.99 |
| American Indian or Alaska Native | 21 (0.55) | 5 (0.54) | 12 (0.51) | 4 (0.74) |  |
| Asian | 323 (8.46) | 90 (9.67) | 189 (8.06) | 44 (8.15) |  |
| Asian, non-Hispanic | 2 (0.05) | 0 (0) | 1 (0.04) | 1 (0.19) |  |
| Black or African American | 124 (3.25) | 32 (3.44) | 71 (3.03) | 21 (3.89) |  |
| Black, non-Hispanic | 2 (0.05) | 1 (0.11) | 1 (0.04) | 0 (0) |  |
| Native American, Hispanic | 0 (0) | 0 (0) | 0 (0) | 0 (0) |  |
| Native American, non-Hispanic | 1 (0.03) | 1 (0.11) | 0 (0) | 0 (0) |  |
| Native Hawaiian or Other Pacific | 15 (0.39) | 4 (0.43) | 8 (0.34) | 3 (0.56) |  |
| Other | 647 (16.95) | 161 (17.29) | 388 (16.54) | 98 (18.15) |  |
| Other, Hispanic | 5 (0.13) | 2 (0.21) | 3 (0.13) | 0 (0) |  |
| Other, non-Hispanic | 0 (0) | 0 (0) | 0 (0) | 0 (0) |  |
| Patient Refused | 121 (3.17) | 37 (3.97) | 66 (2.81) | 18 (3.33) |  |
| Unknown | 134 (3.51) | 42 (4.51) | 77 (3.28) | 15 (2.78) |  |
| White | 2021 (52.95) | 480 (51.56) | 1283 (54.69) | 258 (47.78) |  |
| White, Hispanic | 0 (0) | 0 (0) | 0 (0) | 0 (0) |  |
| White, non-Hispanic | 36 (0.94) | 9 (0.97) | 22 (0.94) | 5 (0.93) |  |
| No response | 365 (9.56) | 67 (7.2) | 225 (9.59) | 73 (13.52) |  |
| **Marital Status** |  |  |  |  | 0.76 |
| Married | 1991 (52.16) | 529 (56.82) | 1230 (52.43) | 232 (42.96) |  |
| Separated | 99 (2.59) | 15 (1.61) | 59 (2.51) | 25 (4.63) |  |
| Widowed | 157 (4.11) | 49 (5.26) | 89 (3.79) | 19 (3.52) |  |
| Never Married | 684 (17.92) | 162 (17.40) | 420 (17.90) | 102 (18.89) |  |
| Living Together | 243 (6.37) | 55 (5.91) | 152 (6.48) | 36 (6.67) |  |
| Divorced | 396 (10.37) | 81 (8.7) | 250 (10.66) | 65 (12.04) |  |
| No response | 247 (6.48) | 40 (4.3) | 146 (6.22) | 61 (11.3) |  |
| **Education (years):** |  |  |  |  | 0.21 |
| ≤12 | 108 (2.83) | 25 (2.69) | 53 (2.26) | 30 (5.56) |  |
| 13-16 | 1140 (29.87) | 241 (25.89) | 700 (29.84) | 199 (36.85) |  |
| 17-20 | 1995 (52.27) | 529 (56.82) | 1245 (53.07) | 221 (40.93) |  |
| ≥21 | 331 (8.67) | 93 (9.99) | 207 (8.82) | 31 (5.74) |  |
| No response | 243 (6.37) | 43 (4.62) | 141 (6.01) | 59 (10.93) |  |
| **Longitudinal dataset (at baseline)** |  |  |  |  |  |
| **Total:** | 1273 (7.70) | 263 (20.66) | 827 (64.96) | 183 (14.38) |  |
| **Age (years):** |  |  |  |  | 0.14 |
| 18-29 | 169 (13.28) | 34 (12.93) | 111 (13.42) | 24 (13.11) |  |
| 30-39 | 184 (14.45) | 35 (13.31) | 123 (14.87) | 26 (14.21) |  |
| 40-49 | 263 (20.66) | 42 (15.97) | 172 (20.8) | 49 (26.78) |  |
| 50-59 | 275 (21.6) | 47 (17.87) | 181 (21.89) | 47 (25.68) |  |
| 60-69 | 229 (17.99) | 56 (21.29) | 144 (17.41) | 29 (15.85) |  |
| ≥70 | 151 (11.86) | 49 (18.63) | 95 (11.49) | 7 (3.83) |  |
| No response | 2 (0.16) | 0 (0) | 1 (0.12) | 1 (0.55) |  |
| **Sex** |  |  |  |  | 0.80 |
| Female | 865 (67.95) | 172 (65.4) | 560 (67.71) | 133 (72.68) |  |
| Male | 367 (28.83) | 84 (31.94) | 239 (28.9) | 44 (24.04) |  |
| No response | 41 (3.22) | 7 (2.66) | 28 (3.39) | 6 (3.28) |  |
| **Ethnicity** |  |  |  |  | 0.89 |
| Hispanic/Latino | 129 (10.13) | 31 (11.79) | 71 (8.59) | 27 (14.75) |  |
| Non-Hispanic/Non-Latino | 998 (78.4) | 207 (78.7) | 652 (78.84) | 139 (75.96) |  |
| Patient refused | 43 (3.38) | 7 (2.66) | 30 (3.63) | 6 (3.28) |  |
| Unknown | 47 (3.69) | 10 (3.8) | 33 (3.99) | 4 (2.19) |  |
| No response | 56 (4.4) | 8 (3.04) | 41 (4.96) | 7 (3.83) |  |
| **Race** |  |  |  |  | 0.95 |
| American Indian or Alaska Native | 10 (0.79) | 3 (1.14) | 6 (0.73) | 1 (0.55) |  |
| Asian | 90 (7.07) | 20 (7.6) | 56 (6.77) | 14 (7.65) |  |
| Asian, non-Hispanic | 0 (0) | 0 (0) | 0 (0) | 0 (0) |  |
| Black or African American | 31 (2.44) | 7 (2.66) | 15 (1.81) | 9 (4.92) |  |
| Black, non-Hispanic | 0 (0) | 0 (0) | 0 (0) | 0 (0) |  |
| Native American, Hispanic | 0 (0) | 0 (0) | 0 (0) | 0 (0) |  |
| Native American, non-Hispanic | 0 (0) | 0 (0) | 0 (0) | 0 (0) |  |
| Native Hawaiian or Other Pacific | 3 (0.24) | 1 (0.38) | 1 (0.12) | 1 (0.55) |  |
| Other | 239 (18.77) | 58 (22.05) | 136 (16.44) | 45 (24.59) |  |
| Other, Hispanic | 0 (0) | 0 (0) | 0 (0) | 0 (0) |  |
| Other, non-Hispanic | 0 (0) | 0 (0) | 0 (0) | 0 (0) |  |
| Patient Refused | 35 (2.75) | 7 (2.66) | 22 (2.66) | 6 (3.28) |  |
| Unknown | 51 (4.01) | 8 (3.04) | 35 (4.23) | 8 (4.37) |  |
| White | 740 (58.13) | 148 (56.27) | 501 (60.58) | 91 (49.73) |  |
| White, Hispanic | 1 (0.08) | 0 (0) | 1 (0.12) | 0 (0) |  |
| White, non-Hispanic | 12 (0.94) | 2 (0.760) | 10 (1.21) | 0 (0) |  |
| No response | 61 (4.79) | 9 (3.42) | 44 (5.32) | 8 (4.37) |  |
| **Marital Status** |  |  |  |  | 0.52 |
| Married | 688 (54.05) | 151 (57.41) | 461 (55.74) | 76 (41.53) |  |
| Separated | 19 (1.49) | 2 (0.76) | 12 (1.45) | 5 (2.73) |  |
| Widowed | 42 (3.3) | 14 (3.32) | 22 (2.66) | 6 (3.28) |  |
| Never Married | 272 (21.37) | 49 (18.63) | 174 (21.04) | 49 (26.78) |  |
| Living Together | 80 (6.28) | 18 (6.84) | 49 (5.93) | 13 (7.1) |  |
| Divorced | 157 (12.33) | 27 (10.27) | 100 (12.09) | 30 (16.39) |  |
| No response | 15 (1.18) | 2 (0.76) | 9 (1.09) | 4 (2.19) |  |
| **Education (years):** |  |  |  |  | 0.06 |
| ≤12 | 21 (1.65) | 4 (1.52) | 12 (1.45) | 5 (2.73) |  |
| 13-16 | 384 (30.16) | 72 (27.37) | 231 (27.93) | 81 (44.26) |  |
| 17-20 | 747 (58.68) | 159 (60.46) | 502 (60.70) | 86 (46.99) |  |
| ≥21 | 109 (8.56) | 23 (8.75) | 77 (9.31) | 9 (4.92) |  |
| No response | 12 (0.94) | 5 (1.9) | 5 (0.6) | 2 (1.09) |  |

### Cluster discovery, characterization, reliability, and validity

The dendrogram reflecting results of the AHCA as implemented on the training dataset is shown in Figure 1B. Based on the gap statistic, we grouped patients into an optimal number of three clusters (Figure 1C). In line with our expectation, the negative affect-related clustering symptoms of depression, anxiety, and anger were the most important factors driving the clustering process, ranked 1^st^, 2^nd^, and 4^th^, respectively, and contributing 42.4% to the overall separability between clusters (Figure 1D).

We labeled the clusters Cluster1, Cluster2, and Cluster3 to reflect the graded scale of severity that characterized all clustering symptoms (Table 2; Figure2A-I), as well as all pain-specific measures (Table 2; Figure 2J-R). These results provided initial validation of these clusters such that: Cluster1 reflects the least severe condition, Cluster3 the worst, and Cluster2 in between, with substantial effect sizes across all comparisons (Table 2). Since PROMIS instruments are normed to the general US population, we could inform that Cluster1 was on average 0.60 standard deviations (SD) better than the norm in the clustering symptoms, but 0.46SD worse than the norm in the subset of PROMIS-based pain-specific measures. Cluster2 was 0.36SD and 1.05SD, and Cluster3 was 1.20SD and 1.54SD, all worse than the norm in the clustering symptoms and pain-specific measures.

**Table 2.** Clustering symptoms and pain-specific measures as per the three clusters in the *training* dataset. M=mean, SD=standard deviation; C1=Cluster1, C2=Cluster2; C3=Cluster3. *Bonferroni threshold for ANOVA main effects of cluster is set at *p*=0.0028 ^Bonferroni threshold for t-test comparisons between each two clusters is set at *p*=0. 0009

|  | Descriptive (M±SD) |  |  | Main Effect of Cluster |  |  | C1 vs. C2 |  | C1 vs. C3 |  | C2 vs. C3 |  |
| --- | --- | --- | --- | --- | --- | --- | --- | --- | --- | --- | --- | --- |
|  | C1 | C2 | C3 | F | df | P* | P^ | Cohen's D | P^ | Cohen's D | P^ | Cohen's D |
| **Clustering symptoms:** |  |  |  |  |  |  |  |  |  |  |  |  |
| Fatigue | 47.36±8.82 | 57.97±7.63 | 67.64±6.68 | 4849.83 | 2, 11445 | <0.0001 | <0.0001 | 1.29 | <0.0001 | 2.59 | <0.0001 | 1.35 |
| Sleep Disturbance | 48.84±8.92 | 55.26±7.35 | 64.24±7.58 | 2742.75 | 2, 11445 | <0.0001 | <0.0001 | 0.79 | <0.0001 | 1.86 | <0.0001 | 1.20 |
| Sleep Impairment | 45.78±9.09 | 55.45±6.79 | 65.92±6.55 | 5218.07 | 2, 11445 | <0.0001 | <0.0001 | 1.21 | <0.0001 | 2.54 | <0.0001 | 1.57 |
| Depression | 42.29±6.82 | 53.87±6.34 | 63.82±6.95 | 7515.97 | 2, 11445 | <0.0001 | <0.0001 | 1.76 | <0.0001 | 3.13 | <0.0001 | 1.50 |
| Anxiety | 44.2±7.26 | 54.76±6.83 | 64.81±6.56 | 6334.6 | 2, 11445 | <0.0001 | <0.0001 | 1.50 | <0.0001 | 2.98 | <0.0001 | 1.50 |
| Anger | 39.11±7.71 | 49.18±7.38 | 59.26±8.23 | 4860.57 | 2, 11445 | <0.0001 | <0.0001 | 1.33 | <0.0001 | 2.53 | <0.0001 | 1.29 |
| Social Isolation | 38.22±6.67 | 47.48±7.29 | 55.61±7.89 | 4034.82 | 2, 11445 | <0.0001 | <0.0001 | 1.33 | <0.0001 | 2.38 | <0.0001 | 1.07 |
| Emotional Support | 56.94±9.23 | 49.99±8.56 | 47.4±8.81 | 933.58 | 2, 11445 | <0.0001 | <0.0001 | 0.78 | <0.0001 | 1.06 | <0.0001 | 0.30 |
| Satisfaction w/ Soc. Roles | 52.91±9.14 | 41.71±7.1 | 35.79±7.6 | 3614.51 | 2, 11445 | <0.0001 | <0.0001 | 1.37 | <0.0001 | 2.04 | <0.0001 | 0.80 |
| **Pain-specific measures:** |  |  |  |  |  |  |  |  |  |  |  |  |
| Pain Intensity | 4.93±2.34 | 5.86±1.98 | 7.01±1.76 | 734.67 | 2, 11445 | <0.0001 | <0.0001 | 0.50 | <0.0001 | 1.49 | <0.0001 | 0.92 |
| # Bodymap Segments | 7.62±8.43 | 11.39±11.39 | 17.99±16.09 | 461.19 | 2, 9916 | <0.0001 | <0.0001 | 0.27 | <0.0001 | 1.29 | <0.0001 | 0.58 |
| Pain Duration | 86.5±119.99 | 95.91±122.92 | 101.8±116.73 | 9.74 | 2, 9477 | <0.0001 | <0.002 | 0.08 | <0.0001 | 0.18 | 0.061 | 0.07 |
| Pain Interference | 57.34±7.97 | 63.52±6.45 | 69.29±5.78 | 2227.08 | 2, 11445 | <0.0001 | <0.0001 | 1.01 | <0.0001 | 2.62 | <0.0001 | 1.41 |
| Pain Behavior | 54.26±6.82 | 58.39±4.39 | 61.53±3.14 | 1575.07 | 2, 11445 | <0.0001 | <0.0001 | 1.08 | <0.0001 | 2.34 | <0.0001 | 1.41 |
| Physical function | 44.13±10.18 | 37.4±8.49 | 32.5±6.95 | 467.67 | 2, 4081 | <0.0001 | <0.0001 | 0.87 | <0.0001 | 1.94 | <0.0001 | 1.00 |
| Phys. Func.-Mobility | 47.69±9.88 | 41.49±9.26 | 36.79±8.13 | 642.56 | 2, 7361 | <0.0001 | <0.0001 | 0.71 | <0.0001 | 1.66 | <0.0001 | 0.82 |
| Phys. Func.-Up. Extrem. | 46.64±9.52 | 40.49±10.3 | 34.71±9.77 | 640.15 | 2, 7361 | <0.0001 | <0.0001 | 0.61 | <0.0001 | 1.64 | <0.0001 | 0.84 |
| Pain Catastrophizing | 12.67±9.88 | 20.99±10.97 | 32.37±11.15 | 2237.26 | 2, 10795 | <0.0001 | <0.0001 | 0.75 | <0.0001 | 2.54 | <0.0001 | 1.44 |

**Fig. 2.**
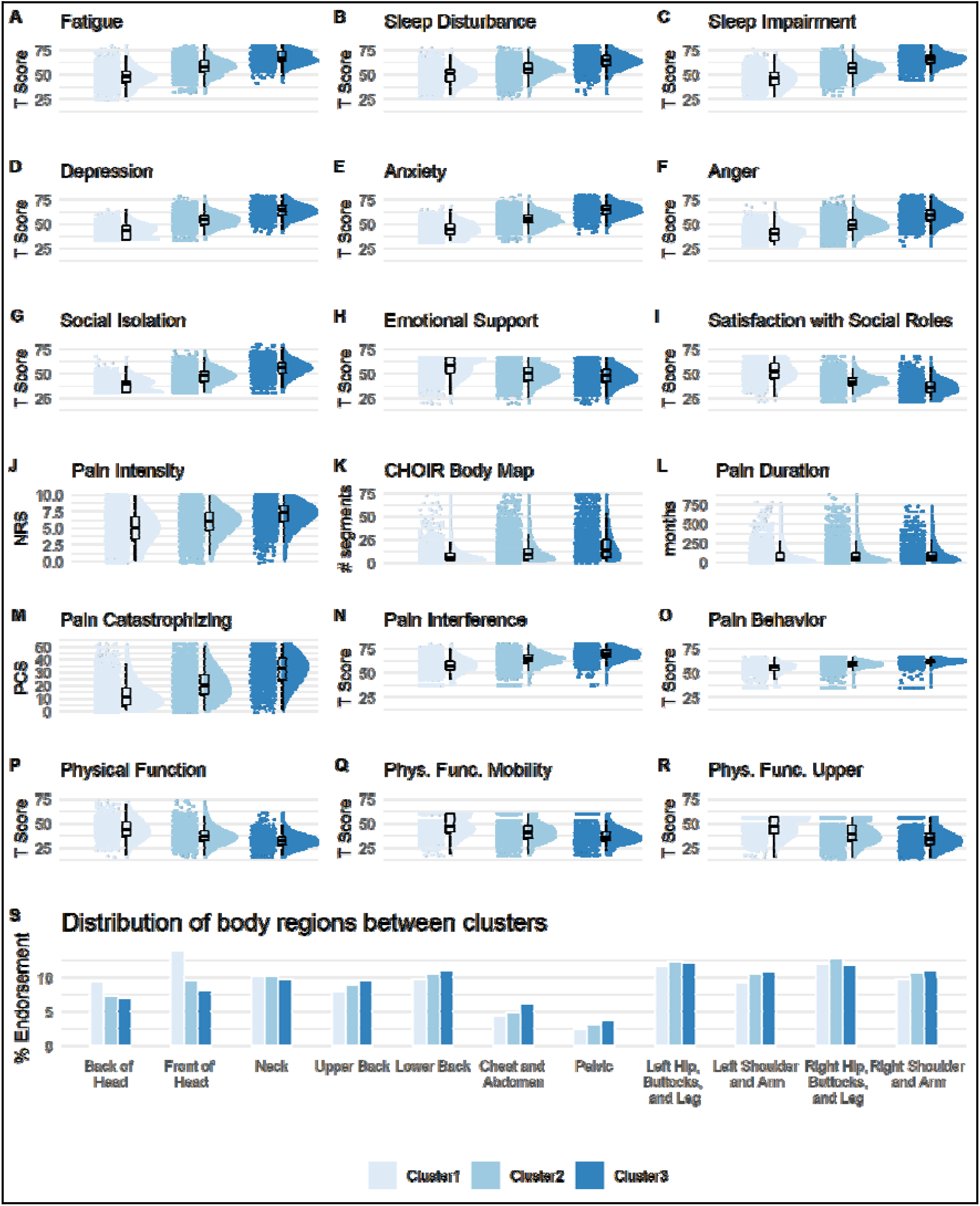
Cluster characterization and diagnostic-like validation. A graded scale of severity is manifested across all clustering symptoms (**A-I**), as well as all pain-specific measures (**J-R**), such that Cluster1 reflects a low severity, Cluster2 a medium severity, and Cluster3 the worst severity. Raincloud plots combining jittered raw data, data distribution, and boxplots were generated using open source code (*83*). Complementary descriptive and inferential statistical information is provided in Table 2. (**S**) The plot shows the % endorsement of 11 body regions as distributed in each of the clusters. There was no significant association in the distribution of % endorsed body regions between the clusters (*p*=0.99). NRS=Numerical Rating Scale; PCS=Pain Catastrophizing Scale.

Although the pattern of severity also manifested in the number of self-reported body segments in pain, with Cluster3 indicative of potential widespread and/or overlapping chronic pain conditions, we found no significant associations between specific body regions (Table S2, Figure S1) and any of the clusters (Chi^2^=3.25, *p*=0.99; Figure2S). Cluster1 was only descriptively associated with more pain in the Front of Head (13.89%) compared to Cluster2 (9.47%) and Cluster3 (8.07%; Chi^2^=1.76, *p*=0.41). Similarly, none of the demographic characteristics were significantly associated with any specific cluster (Table 1). We replicated the same pattern of results across clustering symptoms and pain-specific measures in the *validation* (Table S3, Figure S2) as well as the *longitudinal* datasets (Table S4, Figure S3), except for pain duration, for which we found no differences between the three clusters (*p-values*>0.12). This supported the reliability and validity of the identified clusters. However, two critical questions arose that we addressed in the following two sections.

### Can we obtain a similar clustering solution using only pain intensity?

The clustering solution identified three clusters portraying a graded scale of severity across all clustering symptoms and, importantly, all pain-specific measures. To address whether one variable could be used to obtain a similar clustering solution, we applied AHCA on the training dataset using a common measure for assessing the severity of pain, namely pain intensity. The dendrogram reflecting the results of the AHCA is shown in Figure 3A. The gap statistic indicated an optimal number of one cluster (Figure 3B). We nevertheless selected the non-optimal three-cluster solution to compare it with the clustering symptoms-based solution directly. To visualize this comparison, we applied Principal Components Analysis (PCA) (*23, 24*) on the nine-dimensional clustering symptoms (see Figure S4A for scree plot and Figure S4B for clustering symptoms’ contribution to the first three principal components). To evaluate the separability between the clusters of the two solutions, we plotted the entire training data-set using the first three principal components and colored the data points based on the three clusters of the clustering symptoms solution (Figure 3C) and of the pain intensity solution (Figure 3D). The separability between clusters is clearly seen in the clustering symptoms’ solution, while a substantial overlap is seen in the pain intensity solution, indicating that using pain intensity alone cannot capture a similar solution.

**Fig. 3.**
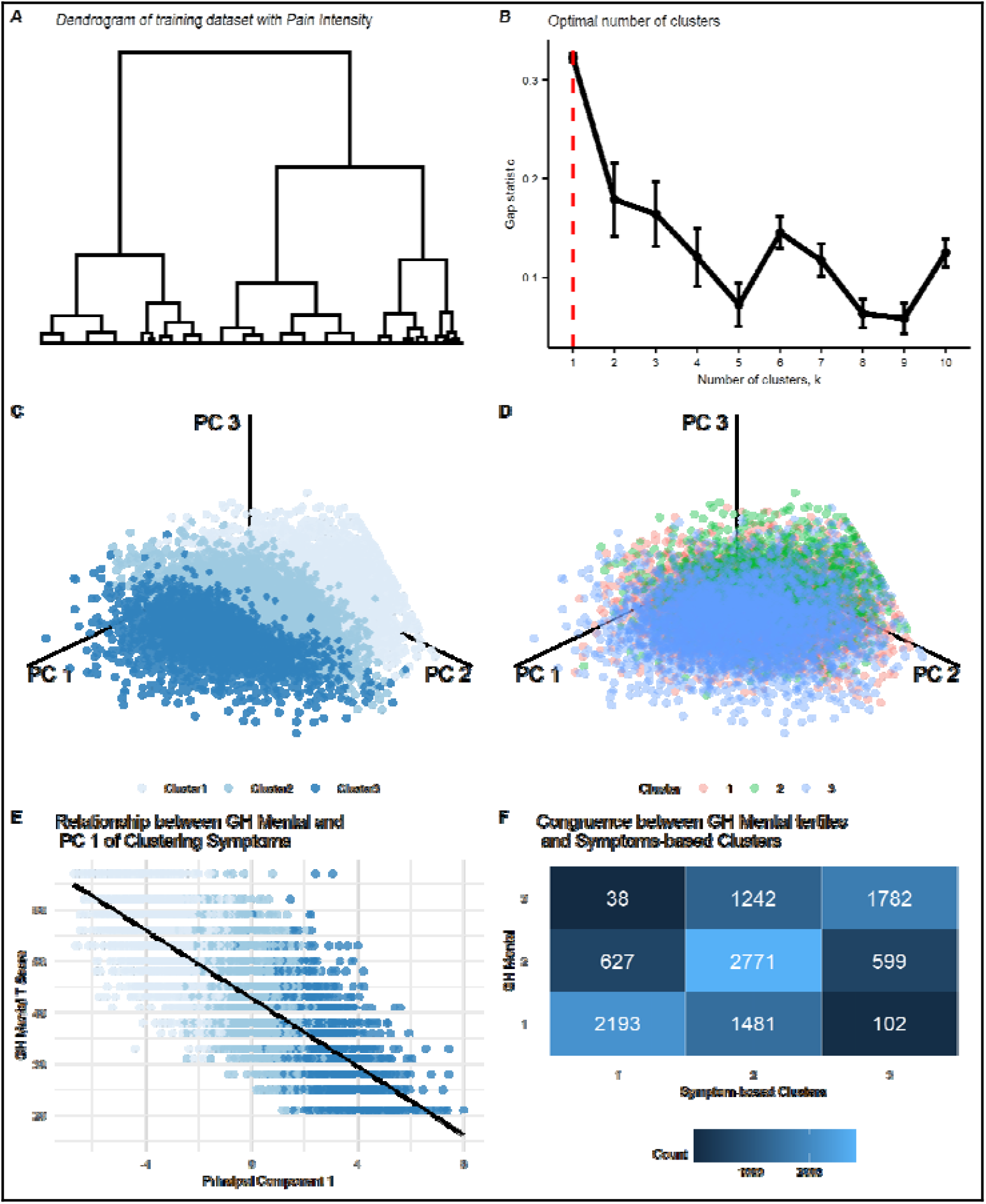
Alternatives to the symptom-based clustering solution. (**A**) The resulting dendrogram reflecting results of the agglomerative hierarchical clustering algorithm (AHCA) as implemented on the training dataset (n=11448) and using the Pain Intensity measure. The tree is not clustered since the optimal solution was of one cluster. (**B**) The plot shows the gap statistic values for different k number of clusters, and indicating with a red dashed line the optimal solution of k=1. The error bars represent one standard error of the estimated gap statistic. (**C**) and (**D**) are plots that show the distribution of all data points in the training dataset on the three primary principal components (PC) derived from the nine-dimensional clustering symptoms, and colored according to either the three clusters generated from the AHCA of these symptoms (**C**) or according to the three clusters generated from the AHCA of pain intensity (**D**). The separability between clusters is clearly seen in the clustering symptoms’ solution, while a substantial overlap is seen in the pain intensity solution. (**E**) The plot shows the correlation between the first PC and the PROMIS global health (GH) mental subscale in the training dataset (available for n=10835): r=-0.78, *p*<0.001. (**F**) Congruence matrix between the clustering symptoms’ three-cluster solution, and the tertiles labeled according to the PROMIS GH mental subscale. The overall level of congruence was 62.26%.

### Does the clustering solution reflect a latent mental health-related construct?

As we initially expected, the negative affect- or mental health-related clustering symptoms were the most important factors driving the clustering process, and contributing most to the first principal component (42.2% of the explained variance; Figure S4B). This may suggest that the underlying structure of the dataset and subsequent clustering solution reflected a mere latent mental health construct. Therefore, by using a measure of mental health we might obtain similar results. To examine this alternative hypothesis, we calculated the Pearson correlation coefficient between the first principal components and the PROMIS Global Health Mental subscale. To note, this measure was available for n=10835 of the training dataset. The first principal component explained 55.42% of the variance in the data structure (Figure S4A), and had a correlation of r=-0.78 with the PROMIS Global Health Mental subscale (Figure 3E). The correlation with the second and third principal components, which explained 12.28% and 8.83% of the variance in the data structure (Figure S4) were r=0.11 and r=-0.02, respectively. As expected, this reconfirms mental health as a key construct in the data’s underlying structure, but not the only.

To further illustrate this point, we split the range of possible PROMIS Global Health Mental scores into tertiles and labeled them in order of severity (1, 2, and 3) to match the clustering symptoms’ cluster solution labeling. We then quantified the level of congruence between these two sets of labels by counting how many patients were assigned by each of the solutions to the same cluster label and how many were mismatched between the clusters (Figure 3F). The level of congruence was 76.73%, 50.44%, and 71.77% for Cluster1, Cluster2, and Cluster3, respectively, and with an overall 62.26% congruence. Together, it is clear that mental health is a primary component in the data’s underlying structure. Still, the proposed clustering solution reflects more than a mere latent mental health-related construct, particularly at the intermediate Cluster2.

### Predictive validation and cluster dynamics over time

After controlling for the time between the two assessments (3-12months), we were able to demonstrate substantial differences between clusters as identified at baseline in all clustering symptoms (Table 3; Figure 4A-I), and pain-specific measures at follow-up (Table 3; Figure 4J-Q). Cluster1 continued to reflect the least severe condition, Cluster3 the worst, and Cluster2 in between, and again with substantial effect sizes across all comparisons (Table 3). These results validate the prognostic-like nature of the clusters and suggest that the graded scale of severity remains consistent at follow-up at the group-level. Nevertheless, cluster identification at follow-up demonstrates that while most patients (n=879, 69.05%) remained within their same cluster between the two time-points, there were movements across clusters (Figure 5A): 180 patients (14.14%) had an improvement in their condition and moved from Cluster3 to Cluster2 (n=69, 5.42%) or to Cluster1 (n=6, 0.47%), and from Cluster2 to Cluster1 (n=105, 8.25%); and 214 patients (16.81%) had a worsening in their condition and moved from Cluster1 to Cluster2 (n=115, 9.03%) or to Cluster3 (n=4, 0.31%), and from Cluster2 to Cluster3 (n=95, 7.46%). We compared the total movement of patients across clusters between time-points (n=394, 30.95%) to a bootstrapped distribution of patients moving across clusters within potential measurement error (M=5.81%±0.54SD; Figure 5B), which indicated a significant amount of movement (t_(df=999)_=1477.18, p<0.0001; Figure 5C). This suggests that the changes across clusters are meaningful, potentially indicating an interaction between treatment effects and regression to the mean (*51*). Importantly, cluster assignment is not a static condition; rather, various factors might impact the long-term dynamics across clusters, offering a window of opportunity for personalized interventions.

**Table 3.** Clustering symptoms and pain-specific measures in the *longitudinal* dataset at follow-up, as per the three clusters assigned at baseline. M=mean, SD=standard deviation; C1=Cluster1, C2=Cluster2; C3=Cluster3. *Bonferroni threshold for ANCOVA main effects of cluster is set at *p*=0.0029 ^Bonferroni threshold for t-test comparisons between each two clusters is set at *p*=0.001

|  | Descriptive (M±SD) |  |  | Main Effect of Cluster |  |  | C1 vs. C2 |  | C1 vs. C3 |  | C2 vs. C3 |  |
| --- | --- | --- | --- | --- | --- | --- | --- | --- | --- | --- | --- | --- |
|  | C1 | C2 | C3 | F | df | p* | p^ | Cohen's D | p^ | Cohen's D | p^ | Cohen's D |
| **Clustering symptoms:** |  |  |  |  |  |  |  |  |  |  |  |  |
| Fatigue | 53.49±10.23 | 59.53±9.33 | 65.75±9.02 | 61.27 | 3, 1269 | <0.0001 | <0.0001 | 0.62 | <0.0001 | 1.27 | <0.0001 | 0.68 |
| Sleep Disturbance | 50.5±9.42 | 56.35±8.62 | 62.76±9.12 | 69.52 | 3, 1269 | <0.0001 | <0.0001 | 0.65 | <0.0001 | 1.32 | <0.0001 | 0.72 |
| Sleep Impairment | 49.68±9.76 | 57.25±8.46 | 64.46±8.72 | 104.78 | 3, 1269 | <0.0001 | <0.0001 | 0.83 | <0.0001 | 1.60 | <0.0001 | 0.84 |
| Depression | 46.19±8.70 | 54.6±8.35 | 63.13±8.39 | 149.00 | 3, 1269 | <0.0001 | <0.0001 | 0.99 | <0.0001 | 1.98 | <0.0001 | 1.02 |
| Anxiety | 47.35±9.04 | 56.33±8.24 | 64.5±8.45 | 153.95 | 3, 1269 | <0.0001 | <0.0001 | 1.04 | <0.0001 | 1.96 | <0.0001 | 0.98 |
| Anger | 42.04±9.74 | 50.36±9.14 | 59.05±9.57 | 121.55 | 3, 1269 | <0.0001 | <0.0001 | 0.88 | <0.0001 | 1.76 | <0.0001 | 0.93 |
| Social Isolation | 40.68±8.25 | 48.54±8.49 | 57.9±8.66 | 149.69 | 3, 1269 | <0.0001 | <0.0001 | 0.94 | <0.0001 | 2.04 | <0.0001 | 1.09 |
| Emotional Support | 54.92±9.64 | 51.66±9.38 | 47.99±8.44 | 20.93 | 3, 1269 | <0.0001 | <0.0001 | 0.34 | <0.0001 | 0.76 | <0.0001 | 0.41 |
| Satisfaction w/ Soc. Roles | 48.37±10.11 | 42.65±8.22 | 36.14±7.51 | 75.65 | 3, 1269 | <0.0001 | <0.0001 | 0.62 | <0.0001 | 1.37 | <0.0001 | 0.83 |
| **Pain-specific measures:** |  |  |  |  |  |  |  |  |  |  |  |  |
| Pain Intensity | 5.12±2.31 | 5.60±2.05 | 6.39±1.93 | 13.76 | 3, 1269 | <0.0001 | <0.005 | 0.24 | <0.0001 | 0.88 | <0.0001 | 0.58 |
| # Bodymap Segments | 8.87±10.84 | 12.44±12.58 | 17.41±15.45 | 14.62 | 3, 1088 | <0.0001 | <0.0005 | 0.25 | <0.0001 | 0.96 | <0.0001 | 0.45 |
| Pain Interference | 60.72±8.29 | 63.63±7.31 | 67.91±7.07 | 33.25 | 3, 1269 | <0.0001 | <0.0001 | 0.40 | <0.0001 | 1.39 | <0.0001 | 0.86 |
| Pain Behavior | 56.23±5.57 | 58.69±4.18 | 61.07±2.65 | 46.04 | 3, 1269 | <0.0001 | <0.0001 | 0.70 | <0.0001 | 1.64 | <0.0001 | 1.27 |
| Physical function | 40.26±9.90 | 36.86±8.39 | 31.96±6.75 | 12.89 | 3, 488 | <0.0001 | <0.005 | 0.45 | <0.0001 | 1.40 | <0.0001 | 1.03 |
| Phys. Func.-Mobility | 43.86±10.51 | 40.04±10.09 | 35.13±9.48 | 18.63 | 3, 777 | <0.0001 | <0.0001 | 0.39 | <0.0001 | 1.22 | <0.0001 | 0.73 |
| Phys. Func.-Up. Extrem. | 44.92±10.00 | 41.69±8.70 | 37.88±8.56 | 14.52 | 3, 777 | <0.0001 | <0.0001 | 0.37 | <0.0001 | 1.14 | <0.0001 | 0.63 |
| Pain Catastrophizing | 12.57±10.99 | 18.72±12.14 | 29.57±12.62 | 73.07 | 3, 1269 | <0.0001 | <0.0001 | 0.50 | <0.0001 | 1.98 | <0.0001 | 1.22 |

**Fig. 4.**
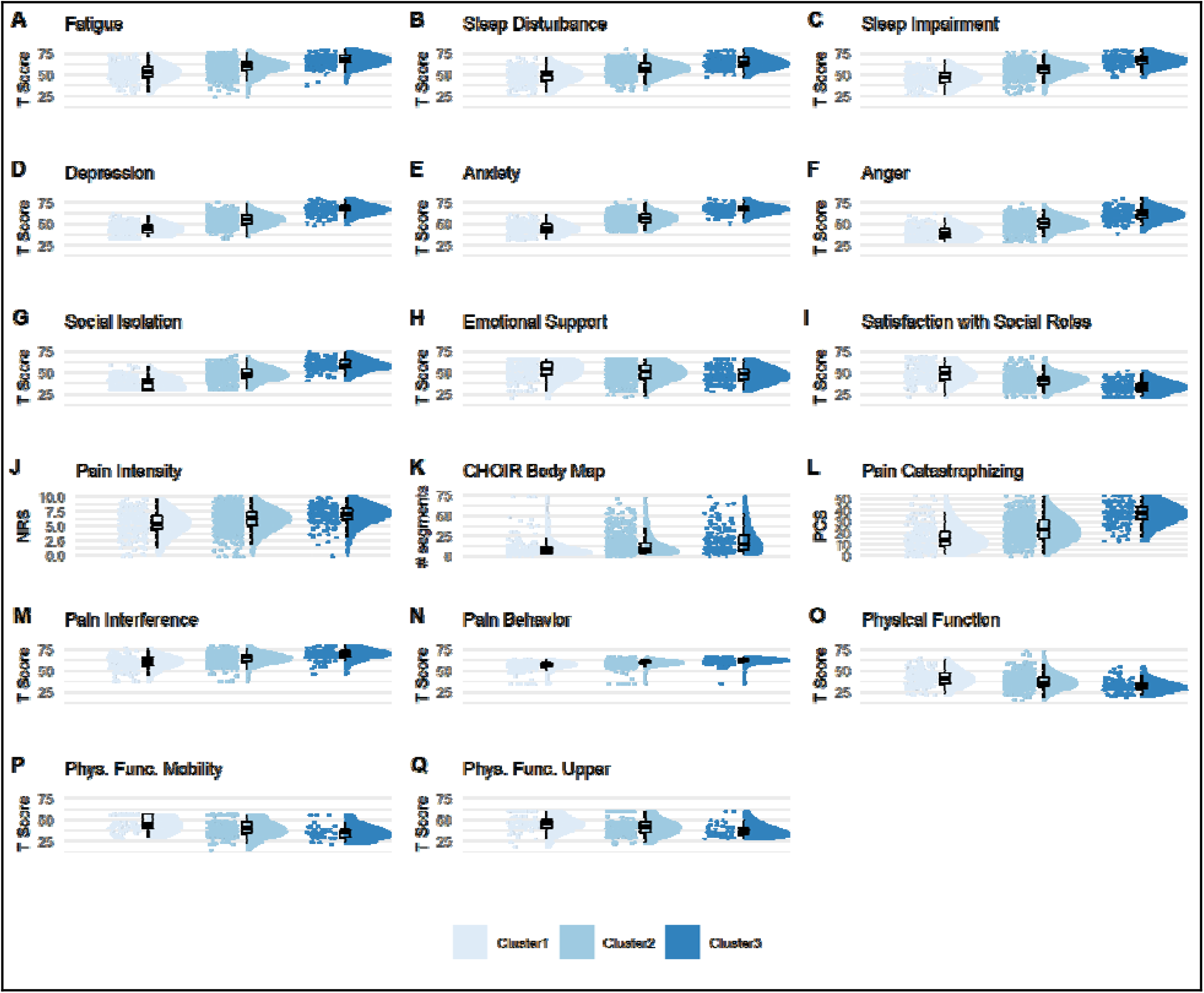
Predictive validation of the clusters. All raincloud plots for the clustering symptoms (**A-I**) as well as for the pain-specific measures (**J-Q**) reflect severity of assessment at follow-up (longitudinal dataset, n=1273), based on cluster identification at baseline. Complementary descriptive and inferential statistical information is provided in Table 3. The graded scale of severity is manifested also here, such that those labeled as Cluster1 at baseline continue to have at the group level the lowest level of severity across all measures, and the same for Cluter2 and Cluster3 being the medium and worst severity, respectively.

**Fig. 5.**
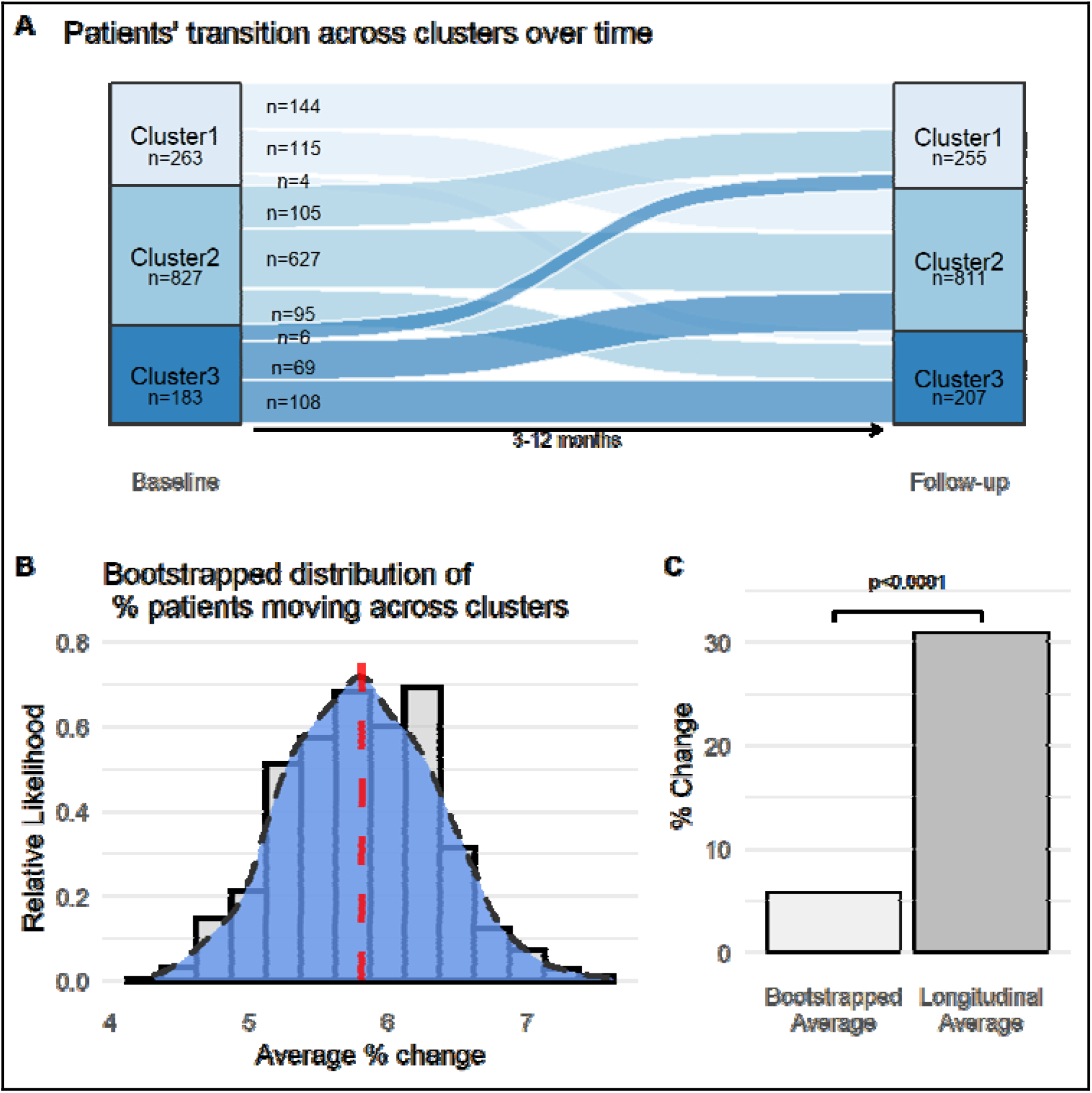
Cluster dynamics over time. (**A**) Sankey plot indicating the transition of patients across clusters over time in the longitudinal dataset (n=1273). Width of lines reflect the extent of movement between time points. One hundred and eighty patients (14.14%) had an improvement in their condition, 214 patients (16.81%) had a worsening in their condition, and 879 (69.05%) remained in the same cluster, between baseline assessment and follow-up. (**B**) Plot of the smoothed kernel density estimate, or relative likelihood, of observing a % change in the bootstrapped distribution of patients moving across clusters within a randomized range of ±3 point measurement error. The red dashed line marks the average set at M=5.81%±0.54SD. The smoothed curve indicates the exact likelihood of observing an exact percent change (i.e., the x-axis value). The bars behind the density curve reflect the same information, averaged at 0.25 sized bins. (**C**) Bar plot comparing the average percent change of patients moving across clusters from the bootstrap distribution (Standard Error=0.017, too small to be seen), with the actual 30.95% found over time in the longitudinal dataset.

## Discussion

In our study we offer a novel biopsychosocial-inspired approach to classify patients with chronic pain, resulting in the identification of three robust idiosyncratic groups of patients, and generating putative markers that can classify current and predict future severity of chronic pain in a graded manner, regardless of their formal diagnosis or their underlying etiology. We applied a data-driven clustering algorithm on multidimensional self-reported symptom assessments that are agnostic to pain. These assessments were collected through CHOIR, Stanford’s registry-based, learning health care system (*30*), and belonging to more than 16 thousand real-world patients seeking treatment at a tertiary academic pain clinic. These findings can be instrumental in supporting treatment selection and pain management in a personalized healthcare platform, especially in the current forward-triage approach to healthcare in which a clinician might not be able to physically examine a patient (*52*). Moreover, findings inspire further research into the biological and behavioral mechanisms that characterize the identified clusters.

The three identified groups reflected a graded scale of severity. They were therefore labeled Cluster1, Cluster2, and Cluster3, with higher numbers indicative of a more severe condition, as shown in all assessments, including those used for clustering as well as those specific for pain, except for pain duration since onset of chronic pain. No apparent demographic factors significantly differed between clusters. The overall group characteristics initially discovered on a subset of more than 11 thousand patients reliably reproduced in two additional subsets consisting of about five thousand patients. The sample size utilized in the development and validation of these clusters (>16K) is consdireably larger compared to even the largest samples (∼5K) used in previous efforts (*11, 12, 14*). Moreover, one of our subsets comprising 1273 patients included follow-up assessments, the severity of which were predicted based on the baseline cluster assignment. Examining the dynamics across clusters between baseline and follow-up assessments indicated that cluster assignment is not a static condition, suggesting that various factors might impact improvement or worsening of the pain condition. Thus, beyond the diagnostic- and prognostic-like nature of these symptom-based putative markers, future clinical and research efforts can examine whether and to what extent they will indicate response to various treatments (*20, 21*).

One of the biggest challenges for chronic pain healthcare is identifying safe and effective treatments tailored to the patient’s particular needs. The symptom-based classification system proposed here addresses this challenge by reversing the common paradigm – it uses the endpoint, as in patient-reported symptoms, to classify the condition of pain and aiming in subsequent research to provide the underlying mechanistic basis. Similar evidence-based approaches have called for chronic pain classification systems that focus on the fine-grained multidimensional and mechanistic substrates of chronic pain conditions (*53, 54*). However, having potentially too many dimensions for classification, and the costs and burdensome nature of medical tests required for such classification processes may make translating them into clinically interpretable and applicable tools challenging (*55*). Subsequently, in most clinical settings, as in research, chronic pain continues to be diagnosed predominantly by the relevant anatomical location of pain (*56*). In contrasts, our computational approach is easily interpretable and substantially cost-effective since it relies on a minimal set of self-reported assessments that can be completed using an electronic device from almost any place, in about 15 minutes, and with hardly any need for assistance from staff. Moreover, unlike the dominant diagnostic system, we find no associations between location of experienced pain and identified cluster. Nevertheless, the number of body regions in pain increased with severity, indicating that patients with widespread and/or overlapping chronic pain conditions suffer more than those with a more localized pain condition. In line with previous research (*12, 57*), findings thus evidence a diminished reliance on specific anatomical locations of experienced pain when assessing and classifying the severity of impairment in primary pain conditions, and potentially when considering treatment avenues.

As expected, negative affect-related symptoms emerged as key factors driving the clustering process. Researchers have previously demonstrated similar negative affect-related metrics to be central in clustering patients with chronic pain (*11, 12*). As we further confirmed, a global measure of mental health was a key construct in the data’s underlying structure. This reverberates with the crucial role of mental health in chronic pain (*9, 28*). Currently, there is an ongoing paradigm shift in psychology and psychiatry, calling for the classification of psychopathology as a hierarchy of continuous dimensions rather than describing it through discrete diagnostic categories (*58*). On top of the hierarchy is a global factor termed the “p factor”, generally ranging from low to high psychopathological severity, and cutting through all psychopathological disorders to account for their nonspecific and overlapping manifestation of symptoms (*59, 60*). The resemblance to chronic pain is astounding. The empirical findings presented here suggest that pain as a field should consider establishing a similar hierarchy of continuous global transdiagnostic dimensions to improve the ability to address the challenges of chronic pain. Moreover, this echoes our contemporary perspective on the need for more synergistic interactions between the research and clinical fields of pain and mental health, specifically regarding the centrality of affective components to these fields (*28*).

Our findings build on existing notions of a general graded scale of severity of chronic pain illness (*57, 61, 62*). However, our approach extends previous efforts in terms of the combination of scale, scope, computational approach, and especially in that we use multidimensional domain-general symptoms that are agnostic to pain. This is advantageous for two main reasons. First, it may highlight potentially modifiable targets for intervention. As we anticipated, negative affect-related factors, namely depression, anxiety, and anger, were key drivers in cluster assignment at the group level. Fortunately, there is a flourishing of treatment strategies aimed to reduce negative affect-related symptomatology (*63*–*68*). Moreover, findings indicate that the distribution of symptoms severity and of patients across clusters is to a certain extent blended (e.g., Figure 2A-R, Figure 3C). This suggests that a healthcare clinician may consider the particular pattern of symptoms at the individual patient level regarding the assigned cluster and utilize this information to guide and support clinical decision-making contextually. For example, we may envision a patient assigned to the lower severity Cluster1, but has relatively high levels of sleep dysfunction that a clinician could address with specific sleep-related treatments (*69*).

A second advantage of the domain-general symptoms approach is that it may be implemented and potentially generalizable to other chronic illnesses requiring symptom management beyond the specific pathophysiology of their disease, like cancer, immune disorders, and cardiovascular diseases among many others. Notably, the graded classification of severity resonates with other illnesses that are characterized by a staged progression of disease, like cancer (*70*), heart (*71*) and kidney diseases (*72*), diabetes (*73*), and more. Here, however, we did not use objective and etiological based metrics, and future integration of genetic, metabolic, inflammatory, and/or anatomical and functional neuroimaging metrics can substantially improve our understanding of the biological mechanisms underlying the identified symptom-based clusters and potentially lead to improved (bio)marker properties (*20, 21*).

The US National Pain Strategy has drawn attention to a sub-group of chronic pain patients – those with persistent high-impact chronic pain. These patients suffer from the most severe and debilitating illness, substantially restricting and interfering with daily life activities, and requiring increased healthcare expenditure (*3, 74, 75*). The prevalence of high-impact chronic pain is estimated to be between 5-15% of the adult US population (10-30 million people) (*3, 5, 75*). Compared to lower but still clinically significant chronic pain, high-impact chronic pain was associated with unfavorable health outcomes, limitations in daily activity, negative coping strategies, elevated distress, increased health care costs, and higher usage and dosage of opioid medication (*62, 76*). With the potential collateral personal, societal, and financial impact of long-term opioid medication (*77*), it is particularly crucial to better identify and understand people suffering from and at increased risk of high-impact chronic pain. Within our proposed classification system, Cluster3 may be reflective of such a group of patients: an overall most severe condition characterizes it, manifested at the group level by highest levels of pain interference, widespread and/or overlapping chronic pain conditions, low levels of physical function, fatigue, depression, and basically in every measure that we assessed. Early identification of these patients is essential for the provision of more comprehensive and costly pain assessments (e.g., psychological, medical, etcetera) that better inform treatment approaches (e.g., physical or psychosocial therapy, medical interventions, etcetera).

There are notable limitations to our study. While our cohort is uniquely large, it is restricted to the San Francisco Bay Area and the outlining Northern California region, and potentially also to patients who can afford specialized medical treatment in a tertiary academic clinic. Future efforts will need to generalize our findings to other locations with different demographic, socio-cultural, and economic characteristics. In this regard, there are known demographic disparities related to pain healthcare (*78, 79*) that were not captured by the identified clusters. This may be attributed to the particular characteristics of the cohort (Table S3), for example being primarily White (53.87%) and with above college level of education (61.92%). However, there are some descriptive trends worth noting. Across datasets (Table S3), Cluster3 was generally characterized by younger age, lower education level, more females, more patients identifying as Hispanic or Latino, less of them identifying as White, and less reporting being married. It is essential to highlight that we can only address most of these factors through a systematic change in healthcare. Additionally, in terms of cohort characteristics, we had no data on formal diagnoses within our cohort. Although findings indicate no association between the identified clusters and anatomical pain location, which is commonly used to support formal definitions of chronic pain conditions, this should still be tested and confirmed. Notably, previous CHOIR studies were able to indicate a multitude of formal diagnoses (*25, 26*), including neuropathic, thoracolumbar, orofacial, visceral, and various musculoskeletal pain conditions, as well as fibromyalgia and complex regional pain syndrome, amongst many others, and these can be assumed to be part of the current cohort. Thus, unlike previous clustering efforts that were restricted to specific chronic pain conditions (*12, 13, 15, 17, 19*), our findings seem to be generalizable at least to varying types of chronic pain conditions.

That the assessments used for clustering are based on NIH’s PROMIS system has its limitations since although they were validated for their psychometric properties (*27, 31*–*36*), they are still based on self-reported assessments and thus prone to potential biases and demand characteristics. Other studies using various clustering approaches have incorporated more objective measurements, with a better characterization of their underlying mechanistic substrates. Most have used various multimodal pain sensory testing that map on to various nociceptive pathways (*12, 13, 15, 16, 18, 19*). While incorporating objective measures with better understanding of their underlying pathophysiology is a clear next step for this research, using the PROMIS system offers substantial advantages. PROMIS-based T scores are normed to the general US population and thus easily comparative across cohorts. PROMIS is also an inexpensive and easily administered system, using short forms or computerized adaptive testing to reduce time and patient burdens, and is already in wide usage in many settings, even beyond chronic pain, thus allowing others to take a similar approach as ours, or to engage with our freely available cluster-classifier (<u>TBA</u>) for additional utilization in clinical and research settings. Moreover, previous findings show associations between various PROMIS measures and potential biomarkers in both pain and non-pain clinical contexts (*80*–*82*). Finally, that the clusters differ in pain-specific measure that are non-PROMIS based, such as pain intensity and pain catastrophizing, solidifies the validity and generalizability beyond PROMIS-based measures.

In conclusion, our symptom-based approach and findings offer significant diagnostic- and prognostic-like utility for a cost-effective, graded severity classification system of patients with chronic pain, potentially generalizable to other chronic illnesses. Our study’s exploratory nature requires further research to reconfirm and generalize the identified clusters in different chronic pain cohorts, as well as experimental and mechanistic studies to uncover their etiological basis. Nevertheless, this system promises to support clinical decision-making, impacting the day-to-day functioning of patients with chronic pain, and encourages investigations into new treatment opportunities oriented towards a precision- and evidence-based approach to relieve the burdens of people suffering from chronic illness, and improve their quality of life. It thus reflects a synergy between theory-driven scientific research, clinical care, and technological advancement that aims to facilitate personalized healthcare by closing on the bedside-to-bench-to-bedside loop.

## Data Availability

All data needed to evaluate the conclusions in the paper are present in the paper. Additional data related to this paper may be requested from the authors.

## Author contributions

Original conceptualization: GG; Contribution to conceptualizations: GG, EMC, KAW, MZ, MCK, SCM; Methodology: GG, EMC, MCK; Software and visualization: EMC, GG; Analyses: GG, EMC; Resources and Supervision: SCM; Writing—original draft: GG; Writing—review & editing: GG, EMC, KAW, MZ, MCK, SCM; All authors discussed the analyses, the results, and their interpretation, and approved the final manuscript.

## Supplementary Materials

Figs. S1 to S4

Tables S1 to S5

**Fig. S1.**
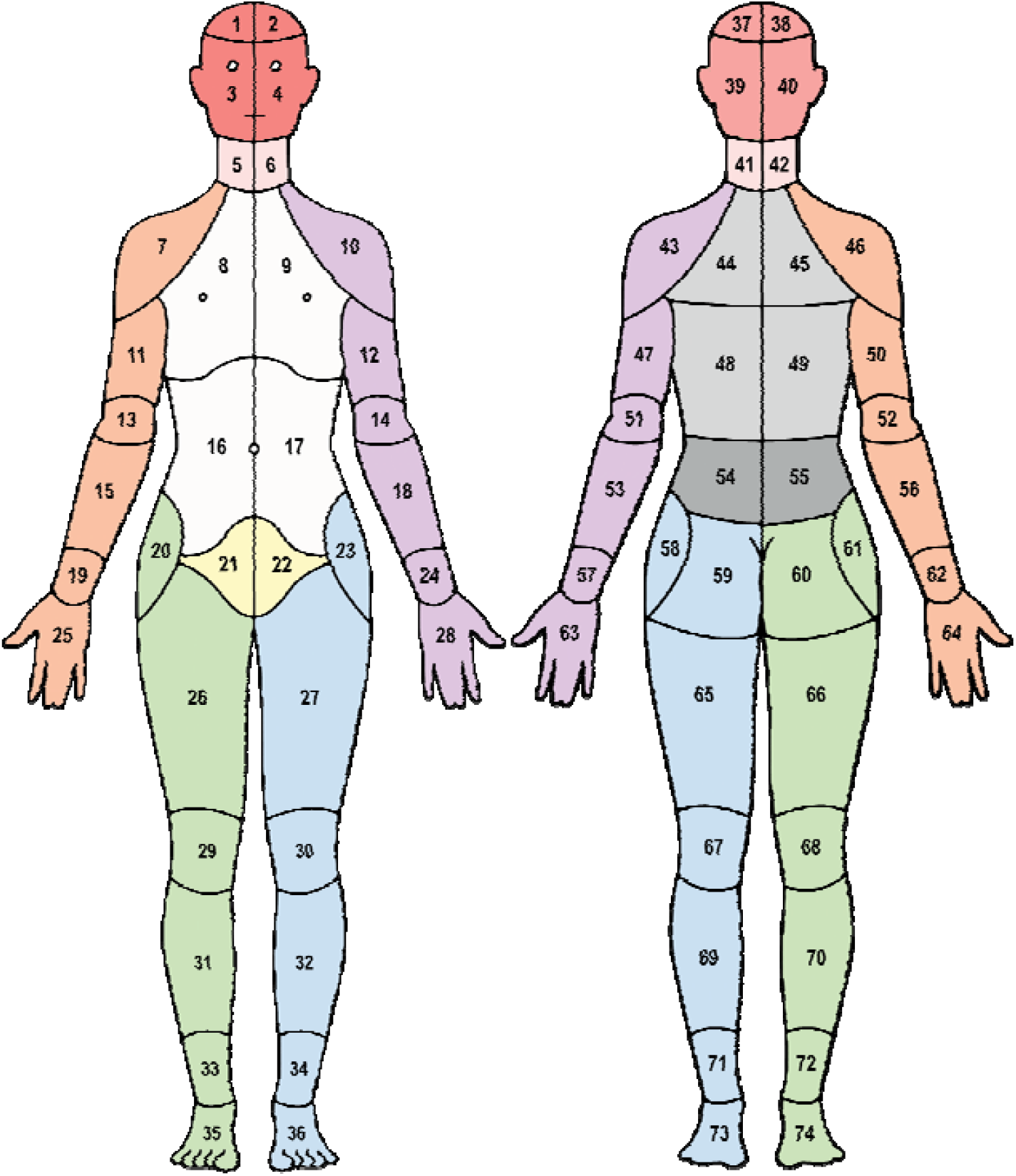
CHOIR body map with 74 numbered segments and 11 colored body regions (see also Table S2).

**Fig. S2.**
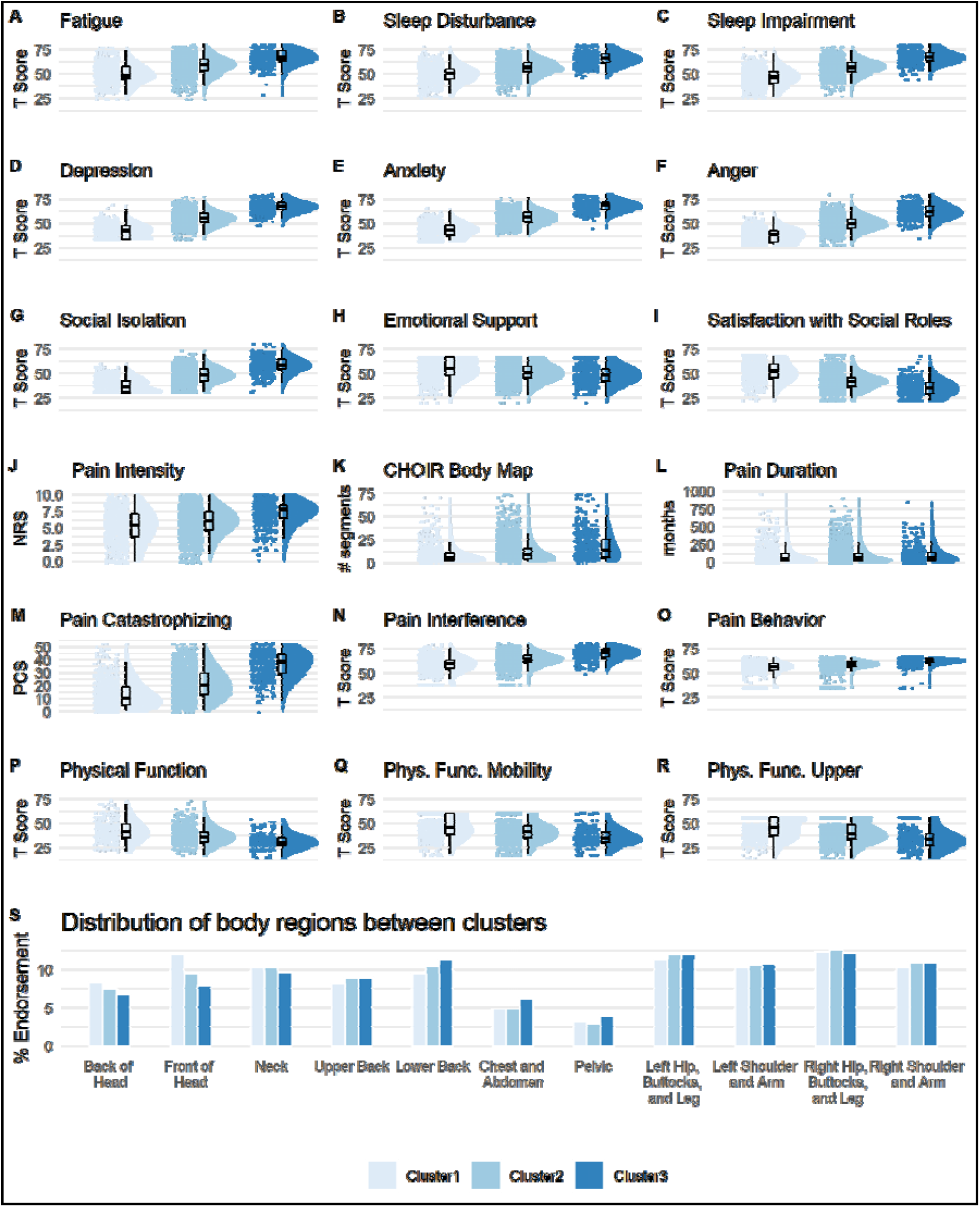
Clusters validation in the *validation* dataset (n=3817). A graded scale of severity is manifested again across all clustering symptoms (A-I), as well as on almost all pain-specific measures (J-R), except for pain duration (L, *p*=0.724), such that Cluster1 reflects a low severity, Cluster2 a medium severity, and Cluster3 the worst severity. Raincloud plots combining jittered raw data, data distribution, and boxplots were generated using open source code (*83*). Complementary descriptive and inferential statistical information is provided in Table S3. (S) The plot shows the % endorsement of 11 body regions as distributed in each of the clusters. There was no significant association in the distribution of % endorsed body regions between the clusters (Chi^2^=1.78, *p*=0.99). NRS=Numerical Rating Scale; PCS=Pain Catastrophizing Scale.

**Fig. S3.**
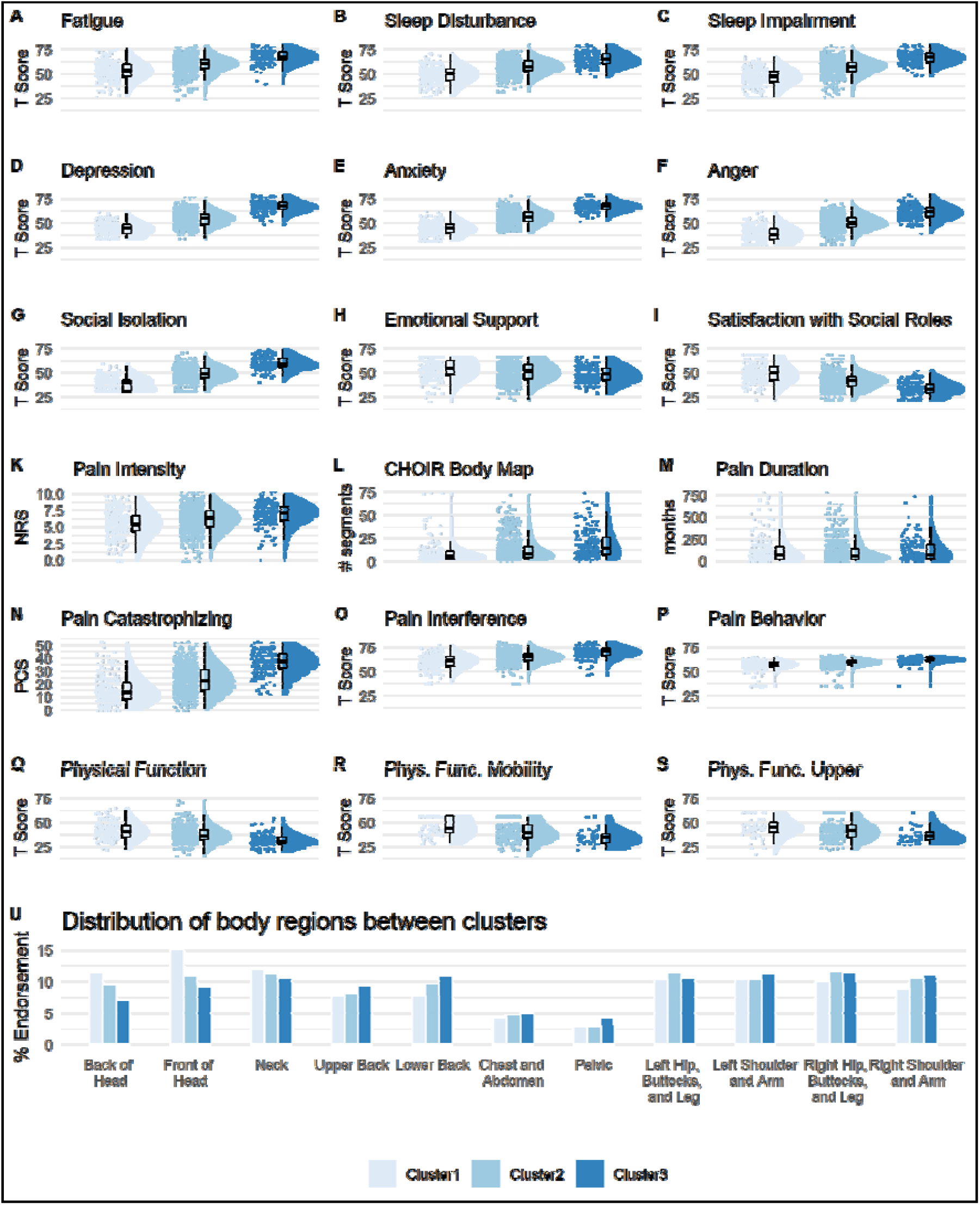
Clusters validation in the *longitudinal* dataset, at baseline (n=1273). A graded scale of severity is manifested again across all clustering symptoms (A-I), as well as on almost all pain-specific measures (J-R), except for pain duration (L, *p*=0.12), such that Cluster1 reflects a low severity, Cluster2 a medium severity, and Cluster3 the worst severity. Raincloud plots combining jittered raw data, data distribution, and boxplots were generated using open source code (*83*). Complementary descriptive and inferential statistical information is provided in Table S4. (S) The plot shows the % endorsement of 11 body regions as distributed in each of the clusters. There was no significant association in the distribution of % endorsed body regions between the clusters (Chi^2^=4.43, *p*=0.99). NRS=Numerical Rating Scale; PCS=Pain Catastrophizing Scale.

**Fig. S4.**
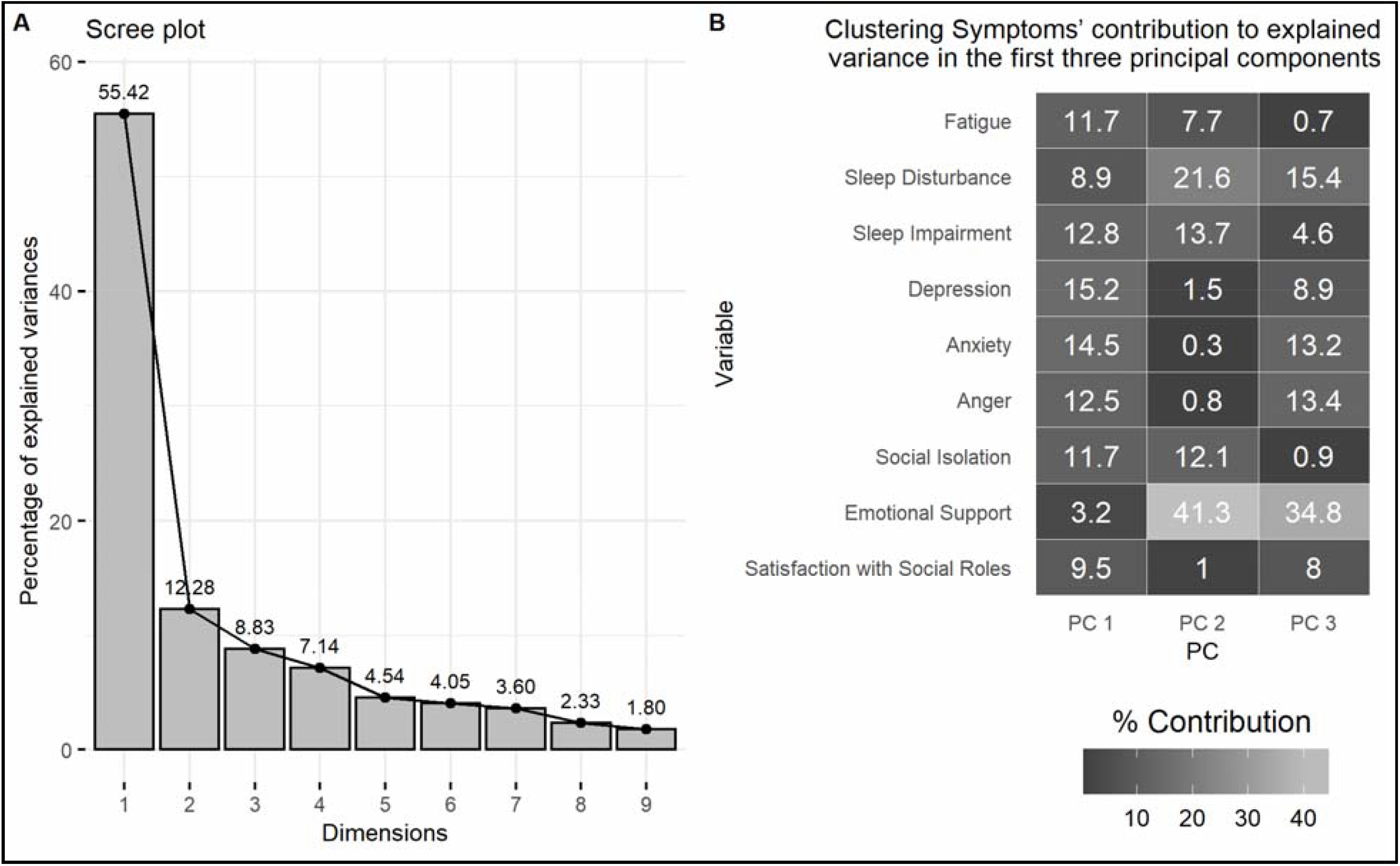
Principal Component Analysis (PCA) applied on the nine clustering symptoms. (**A**) Scree plot indicating the percent of explained variance in the data by each of the nine principal components (PCs). For example, 55.42% of the variance is explained by PC 1. (**B**) The percent contribution of each of the nine clustering symptoms in accounting for the variability in the first three PCs. Variables that are mostly contributing to PC 1 are the most important in explaining the variability in the dataset. The negative affect-related factors Depression, Anxiety, and Anger, are ranked 1^st^, 2^nd^, and 4^th^, respectively, and contribute together 42.2% to PC 1.

**Table S1.** Participants’ demographic information for the entire dataset and across the three clusters. Number of patients is indicated, with % in parenthesis. * Reflects the results of a Chi^2^ test (categories with less than a minimum of 5 patients per group were removed) comparing across clusters.

| N (%) | Total | Cluster1 | Cluster2 | Cluster3 | p* |
| --- | --- | --- | --- | --- | --- |
| **Total:** | 16538 | 4137 (25.02) | 8975 (54.27) | 3426 (20.72) |  |
| **Age (years):** |  |  |  |  | 0.97 |
| 18-29 | 2057 (12.44) | 525 (12.69) | 1070 (11.92) | 462 (13.49) |  |
| 30-39 | 2794 (16.89) | 701 (16.94) | 1463 (16.3) | 630 (18.39) |  |
| 40-49 | 3103 (18.76) | 692 (16.73) | 1640 (18.27) | 771 (22.5) |  |
| 50-59 | 3497 (21.15) | 776 (18.76) | 1930 (21.5) | 791 (23.09) |  |
| 60-69 | 2882 (17.43) | 797 (19.27) | 1606 (17.89) | 479 (13.98) |  |
| ≥70 | 2173 (13.14) | 638 (15.42) | 1247 (13.89) | 288 (8.41) |  |
| No response | 32 (0.19) | 8 (0.19) | 19 (0.21) | 5 (0.15) |  |
| **Sex** |  |  |  |  | 0.96 |
| Female | 10701 (64.71) | 2578 (62.32) | 5824 (64.89) | 2299 (67.1) |  |
| Male | 5283 (31.94) | 1431 (34.59) | 2841 (31.65) | 1011 (29.51) |  |
| No response | 554 (3.35) | 128 (3.09) | 310 (3.45) | 116 (3.39) |  |
| **Ethnicity** |  |  |  |  | 0.97 |
| Hispanic/Latino | 1677 (10.14) | 429 (10.37) | 807 (8.99) | 441 (12.87) |  |
| Non-Hispanic/Non-Latino | 12322 (74.51) | 3125 (75.54) | 6800 (75.77) | 2397 (69.96) |  |
| Patient refused | 514 (3.11) | 137 (3.31) | 278 (3.1) | 99 (2.89) |  |
| Unknown | 611 (3.69) | 168 (4.06) | 331 (3.69) | 112 (3.27) |  |
| No response | 1414 (8.55) | 278 (6.72) | 759 (8.46) | 377 (11) |  |
| **Race** |  |  |  |  | 0.99 |
| American Indian or Alaska Native | 79 (0.48) | 17 (0.41) | 40 (0.45) | 22 (0.64) |  |
| Asian | 1348 (8.15) | 397 (9.6) | 714 (7.96) | 237 (6.92) |  |
| Asian, non-Hispanic | 10 (0.06) | 1 (0.02) | 6 (0.07) | 3 (0.09) |  |
| Black or African American | 560 (3.39) | 141 (3.41) | 270 (3.01) | 149 (4.35) |  |
| Black, non-Hispanic | 10 (0.06) | 4 (0.1) | 3 (0.03) | 3 (0.09) |  |
| Native American, Hispanic | 1 (0.01) | 0 (0) | 0 (0) | 1 (0.03) |  |
| Native American, non-Hispanic | 2 (0.01) | 1 (0.02) | 1 (0.01) | 0 (0) |  |
| Native Hawaiian or Other Pacific | 83 (0.5) | 19 (0.46) | 44 (0.49) | 20 (0.58) |  |
| Other | 2807 (16.97) | 706 (17.07) | 1453 (16.19) | 648 (18.91) |  |
| Other, Hispanic | 16 (0.1) | 3 (0.07) | 12 (0.13) | 1 (0.03) |  |
| Other, non-Hispanic | 8 (0.05) | 2 (0.05) | 5 (0.06) | 1 (0.03) |  |
| Patient Refused | 482 (2.91) | 128 (3.09) | 258 (2.87) | 96 (2.8) |  |
| Unknown | 628 (3.8) | 163 (3.94) | 342 (3.81) | 123 (3.59) |  |
| White | 8909 (53.87) | 2226 (53.81) | 4970 (55.38) | 1713 (50) |  |
| White, Hispanic | 3 (0.02) | 1 (0.02) | 1 (0.01) | 1 (0.03) |  |
| White, non-Hispanic | 154 (0.93) | 41 (0.99) | 86 (0.96) | 27 (0.79) |  |
| No response | 1438 (8.7) | 287 (6.94) | 770 (8.58) | 381 (11.12) |  |
| **Marital Status** |  |  |  |  | 0.72 |
| Married | 8648 (52.29) | 2489 (60.16) | 4722 (52.61) | 1437 (41.91) |  |
| Separated | 355 (2.15) | 54 (1.31) | 182 (2.03) | 119 (3.47) |  |
| Widowed | 636 (3.85) | 162 (3.92) | 357 (3.98) | 117 (3.42) |  |
| Never Married | 3105 (18.77) | 700 (16.92) | 1659 (18.48) | 746 (21.77) |  |
| Living Together | 994 (6.01) | 230 (5.56) | 546 (6.08) | 218 (6.36) |  |
| Divorced | 1771 (10.71) | 337 (8.15) | 973 (10.84) | 461 (13.46) |  |
| No response | 1029 (6.22) | 165 (3.99) | 536 (5.97) | 328 (9.57) |  |
| **Education (years):** |  |  |  |  | 0.36 |
| ≤12 | 471 (2.85) | 102 (2.47) | 224 (2.5) | 145 (4.23) |  |
| 13-16 | 4870 (29.45) | 1055 (25.5) | 2537 (28.27) | 1278 (37.3) |  |
| 17-20 | 8775 (53.06) | 2375 (57.41) | 4882 (54.4) | 1518 (44.31) |  |
| ≥21 | 1465 (8.86) | 443 (10.71) | 838 (9.34) | 184 (5.37) |  |
| No response | 957 (5.79) | 162 (3.92) | 494 (5.5) | 301 (8.79) |  |

**Table S2.** CHOIR body map regions and their associated segments (see also Fig. S1).

| Body region | CHOIR body map segments |
| --- | --- |
| Front of Head | 1, 2, 3, 4 |
| Back of Head | 37, 38, 39, 40 |
| Neck | 5, 6, 41, 42 |
| Chest/Abdomen | 8, 9, 16, 17 |
| Upper Back | 44, 45, 48, 49 |
| Lower Back | 54, 55 |
| Pelvic | 21, 22 |
| Right Shoulder/Arm | 7, 11, 13, 15, 19, 25, 46, 50, 52, 56, 62, 64 |
| Left Shoulder/Arm | 10, 12, 14, 18, 24, 28, 43, 47, 51, 53, 57, 63 |
| Right Hip/Buttocks/Leg | 20, 26, 29, 31, 33, 35, 60, 61, 66, 68, 70, 72, 74 |
| Left Hip/Buttocks/Leg | 23, 27, 30, 32, 34, 36, 58, 59, 65, 67, 69, 71, 73 |

**Table S3.** Clustering symptoms and pain-specific measures as per the three clusters in the *validation* dataset (n=3817). M=mean, SD=standard deviation; C1=Cluster1, C2=Cluster2; C3=Cluster3. *Bonferroni threshold for ANOVA main effects of cluster is set at *p*=0.0028 ^Bonferroni threshold for t-test comparisons between each two clusters is set at *p*=0.0009

|  | Descriptive (M±SD) |  |  | Main Effect of Cluster |  |  | C1 vs. C2 |  | C1 vs. C3 |  | C2 vs. C3 |  |
| --- | --- | --- | --- | --- | --- | --- | --- | --- | --- | --- | --- | --- |
|  | C1 | C2 | C3 | F | df | P* | P^ | Cohen's D | P^ | Cohen's D | P^ | Cohen's D |
| **Clustering symptoms:** |  |  |  |  |  |  |  |  |  |  |  |  |
| Fatigue | 49.84±9.68 | 58.35±8.96 | 67.37±7.61 | 676.62 | 2, 3814 | <0.0001 | <0.0001 | 0.91 | <0.0001 | 2.01 | <0.0001 | 1.09 |
| Sleep Disturbance | 49.13±8.67 | 56.45±7.29 | 65.97±7.54 | 830.78 | 2, 3814 | <0.0001 | <0.0001 | 0.91 | <0.0001 | 2.07 | <0.0001 | 1.28 |
| Sleep Impairment | 46.13±9.1 | 56.56±7.28 | 67.33±6.86 | 1344.73 | 2, 3814 | <0.0001 | <0.0001 | 1.27 | <0.0001 | 2.63 | <0.0001 | 1.52 |
| Depression | 41.77±6.53 | 54.84±6.59 | 67.23±5.93 | 2785.15 | 2, 3814 | <0.0001 | <0.0001 | 1.99 | <0.0001 | 4.08 | <0.0001 | 1.98 |
| Anxiety | 43.14±6.75 | 55.98±6.37 | 68.12±5.68 | 2773.84 | 2, 3814 | <0.0001 | <0.0001 | 1.96 | <0.0001 | 4.00 | <0.0001 | 2.01 |
| Anger | 37.58±6.96 | 50.05±7.03 | 62.33±7.65 | 2175.22 | 2, 3814 | <0.0001 | <0.0001 | 1.78 | <0.0001 | 3.38 | <0.0001 | 1.67 |
| Social Isolation | 37.25±6.65 | 47.94±7.36 | 58.16±7.57 | 1511.44 | 2, 3814 | <0.0001 | <0.0001 | 1.52 | <0.0001 | 2.93 | <0.0001 | 1.37 |
| Emotional Support | 53.99±10.11 | 51.04±8.98 | 48.68±8.84 | 61.47 | 2, 3814 | <0.0001 | <0.0001 | 0.31 | <0.0001 | 0.56 | <0.0001 | 0.26 |
| Satisfaction w/ Soc. Roles | 51.93±10.04 | 41.79±8.17 | 34.73±8.14 | 762.22 | 2, 3814 | <0.0001 | <0.0001 | 1.11 | <0.0001 | 1.88 | <0.0001 | 0.87 |
| **Pain-specific measures:** |  |  |  |  |  |  |  |  |  |  |  |  |
| Pain Intensity | 5.13±2.43 | 5.89±1.96 | 7.21±1.79 | 173.39 | 2, 3814 | <0.0001 | <0.0001 | 0.40 | <0.0001 | 1.50 | <0.0001 | 1.04 |
| # Bodymap Segments | 8.46±9.68 | 11.69±11.39 | 17.71±15.67 | 88.49 | 2, 3279 | <0.0001 | <0.0001 | 0.24 | <0.0001 | 1.15 | <0.0001 | 0.54 |
| Pain Duration | 95.14±132.12 | 97.03±124.25 | 99.51±119.86 | 0.17 | 2, 3613 | 0.846 | 0.724 | 0.02 | 0.575 | 0.05 | 0.71 | 0.03 |
| Pain Interference | 58±8.34 | 63.79±6.56 | 70.39±5.87 | 557.71 | 2, 3814 | <0.0001 | <0.0001 | 0.93 | <0.0001 | 2.67 | <0.0001 | 1.59 |
| Pain Behavior | 54.89±6.73 | 58.65±4.38 | 61.92±3.32 | 371.14 | 2, 3814 | <0.0001 | <0.0001 | 0.97 | <0.0001 | 2.27 | <0.0001 | 1.39 |
| Physical function | 42.62±10.71 | 37.26±8.49 | 31.44±6.92 | 106.97 | 2, 1378 | <0.0001 | <0.0001 | 0.69 | <0.0001 | 1.86 | <0.0001 | 1.19 |
| Phys. Func.-Mobility | 46.09±10.63 | 41.75±9.49 | 36.17±8.42 | 111.29 | 2, 2433 | <0.0001 | <0.0001 | 0.48 | <0.0001 | 1.48 | <0.0001 | 0.94 |
| Phys. Func.-Up. Extrem. | 45.28±10.28 | 40.79±10.05 | 34±10.23 | 128.48 | 2, 2433 | <0.0001 | <0.0001 | 0.44 | <0.0001 | 1.59 | <0.0001 | 0.94 |
| Pain Catastrophizing | 12.86±10.19 | 21.6±11.12 | 36.37±9.91 | 753.77 | 2, 3598 | <0.0001 | <0.0001 | 0.83 | <0.0001 | 2.99 | <0.0001 | 2.11 |

**Table S4.** Clustering symptoms and pain-specific measures as per the three clusters in the *longitudinal* dataset, at baseline (n=1273). M=mean, SD=standard deviation; C1=Cluster1, C2=Cluster2; C3=Cluster3. *Bonferroni threshold for ANOVA main effects of cluster is set at *p*=0.0028 ^Bonferroni threshold for t-test comparisons between each two clusters is set at *p*=0.0009

|  | Descriptive (M±SD) |  |  | Main Effect of Cluster |  |  | C1 vs. C2 |  | C1 vs. C3 |  | C2 vs. C3 |  |
| --- | --- | --- | --- | --- | --- | --- | --- | --- | --- | --- | --- | --- |
|  | C1 | C2 | C3 | F | df | P* | P^ | Cohen's D | P^ | Cohen's D | P^ | Cohen's D |
| **Clustering symptoms:** |  |  |  |  |  |  |  |  |  |  |  |  |
| Fatigue | 53.02±9.47 | 59.37±8.6 | 67.62±7.16 | 155.85 | 2, 1270 | <0.0001 | <0.0001 | 0.70 | <0.0001 | 1.74 | <0.0001 | 1.04 |
| Sleep Disturbance | 48.68±8.87 | 56.85±7.23 | 64.93±7.2 | 252.70 | 2, 1270 | <0.0001 | <0.0001 | 1.01 | <0.0001 | 2.01 | <0.0001 | 1.12 |
| Sleep Impairment | 46.29±8.44 | 56.73±7.85 | 66.78±6.56 | 381.60 | 2, 1270 | <0.0001 | <0.0001 | 1.28 | <0.0001 | 2.71 | <0.0001 | 1.39 |
| Depression | 43.3±6.53 | 54.84±6.65 | 66.97±5.94 | 721.08 | 2, 1270 | <0.0001 | <0.0001 | 1.75 | <0.0001 | 3.79 | <0.0001 | 1.92 |
| Anxiety | 44.26±6.5 | 55.92±6.52 | 67.4±5.48 | 727.29 | 2, 1270 | <0.0001 | <0.0001 | 1.79 | <0.0001 | 3.85 | <0.0001 | 1.91 |
| Anger | 38.79±7.66 | 50.44±7.26 | 61.28±7.13 | 523.95 | 2, 1270 | <0.0001 | <0.0001 | 1.56 | <0.0001 | 3.04 | <0.0001 | 1.51 |
| Social Isolation | 38.24±6.84 | 48.5±7.13 | 59.19±6.44 | 495.58 | 2, 1270 | <0.0001 | <0.0001 | 1.47 | <0.0001 | 3.15 | <0.0001 | 1.57 |
| Emotional Support | 53.43±9.96 | 50.97±9.47 | 47.74±8.17 | 19.79 | 2, 1270 | <0.0001 | <0.0005 | 0.25 | <0.0001 | 0.62 | <0.0001 | 0.37 |
| Satisfaction w/ Soc. Roles | 49.6±9.91 | 41.02±7.81 | 33.52±6.81 | 218.01 | 2, 1270 | <0.0001 | <0.0001 | 0.96 | <0.0001 | 1.89 | <0.0001 | 1.02 |
| **Pain-specific measures:** |  |  |  |  |  |  |  |  |  |  |  |  |
| Pain Intensity | 5.39±2.06 | 6.05±1.85 | 6.95±1.64 | 37.59 | 2, 1270 | <0.0001 | <0.0001 | 0.38 | <0.0001 | 1.19 | <0.0001 | 0.78 |
| # Bodymap Segments | 9.27±11.63 | 12.77±12.44 | 18.88±16.03 | 24.12 | 2, 980 | <0.0001 | 0.0005 | 0.24 | <0.0001 | 1.09 | <0.0001 | 0.54 |
| Pain Duration | 118.85±161.08 | 99.14±123.74 | 114.22±128.99 | 2.10 | 2, 1017 | 0.12 | 0.06 | 0.16 | 0.78 | 0.05 | 0.19 | 0.17 |
| Pain Interference | 59.9±7.02 | 64.73±5.97 | 69.81±5.92 | 140.15 | 2, 1270 | <0.0001 | <0.0001 | 0.81 | <0.0001 | 2.35 | <0.0001 | 1.21 |
| Pain Behavior | 55.86±5.48 | 59.09±3.61 | 61.83±3.07 | 125.90 | 2, 1270 | <0.0001 | <0.0001 | 0.96 | <0.0001 | 2.34 | <0.0001 | 1.26 |
| Physical function | 41.42±8.82 | 37.23±8.16 | 31.74±6.33 | 58.40 | 2, 931 | <0.0001 | <0.0001 | 0.57 | <0.0001 | 1.68 | <0.0001 | 1.23 |
| Phys. Func.-Mobility | 45.1±9.1 | 40.41±9.53 | 34.98±8.36 | 16.21 | 2, 336 | <0.0001 | <0.0005 | 0.52 | <0.0001 | 1.50 | <0.001 | 0.92 |
| Phys. Func.-Up. Extrem. | 45.4±9.53 | 41.84±9.25 | 37.12±8.08 | 10.98 | 2, 336 | <0.0001 | <0.005 | 0.41 | <0.0001 | 1.27 | <0.005 | 0.83 |
| Pain Catastrophizing | 15.27±10.53 | 23.24±10.68 | 36.58±8.85 | 227.25 | 2, 1270 | <0.0001 | <0.0001 | 0.81 | <0.0001 | 2.82 | <0.0001 | 2.13 |

**Table S5.** Agglomerative coefficient values of various linkage methods that were combined with the Euclidian distance metric.

| Linkage methods | Agglomerative coefficient |
| --- | --- |
| Single | 0.440 |
| Average | 0.670 |
| McQuitty | 0.749 |
| Complete | 0.846 |
| **Ward** | **0.920** |

